# An atlas connecting shared genetic architecture of human diseases and molecular phenotypes provides insight into COVID-19 susceptibility

**DOI:** 10.1101/2020.12.20.20248572

**Authors:** Liuyang Wang, Thomas J. Balmat, Alejandro L. Antonia, Florica J. Constantine, Ricardo Henao, Thomas W. Burke, Andy Ingham, Micah T. McClain, Ephraim L. Tsalik, Emily R. Ko, Geoffrey S. Ginsburg, Mark R. DeLong, Xiling Shen, Christopher W. Woods, Elizabeth R. Hauser, Dennis C. Ko

## Abstract

While genome-wide associations studies (GWAS) have successfully elucidated the genetic architecture of complex human traits and diseases, understanding mechanisms that lead from genetic variation to pathophysiology remains an important challenge. Methods are needed to systematically bridge this crucial gap to facilitate experimental testing of hypotheses and translation to clinical utility. Here, we leveraged cross-phenotype associations to identify traits with shared genetic architecture, using linkage disequilibrium (LD) information to accurately capture shared SNPs by proxy, and calculate significance of enrichment. This shared genetic architecture was examined across differing biological scales through incorporating data from catalogs of clinical, cellular, and molecular GWAS. We have created an interactive web database (interactive Cross-Phenotype Analysis of GWAS database (iCPAGdb); http://cpag.oit.duke.edu) to facilitate exploration and allow rapid analysis of user-uploaded GWAS summary statistics. This database revealed well-known relationships among phenotypes, as well as the generation of novel hypotheses to explain the pathophysiology of common diseases. Application of iCPAGdb to a recent GWAS of severe COVID-19 demonstrated unexpected overlap of GWAS signals between COVID-19 and human diseases, including with idiopathic pulmonary fibrosis driven by the *DPP9* locus. Transcriptomics from peripheral blood of COVID-19 patients demonstrated that *DPP9* was induced in SARS-CoV-2 compared to healthy controls or those with bacterial infection. Further investigation of cross-phenotype SNPs with severe COVID-19 demonstrated colocalization of the GWAS signal of the *ABO* locus with plasma protein levels of a reported receptor of SARS-CoV-2, CD209 (DC-SIGN), pointing to a possible mechanism whereby glycosylation of CD209 by *ABO* may regulate COVID-19 disease severity. Thus, connecting genetically related traits across phenotypic scales links human diseases to molecular and cellular measurements that can reveal mechanisms and lead to novel biomarkers and therapeutic approaches.

## Introduction

Genome-wide association studies (GWAS) have identified hundreds of thousands of genomic regions that are associated with complex human traits and have increased our understanding of the genetic architecture of human disease (Visscher et al., 2017). While GWAS now utilize even millions of subjects through leveraging electronic medical record data (Bycroft et al., 2018; McCarty et al., 2011), progress towards understanding how identified genetic variants alter cellular function and physiology remains elusive. More efficient mechanisms are needed for translating knowledge of genetic disease risk and severity into insight of the underlying physiology. Integrating analysis of GWAS across different scales of biological phenotypes (molecular, cellular, and organismal) may provide novel insight into how genetic variants influence complex traits.

Comparative analyses of GWAS have revealed that numerous, seemingly unrelated traits are connected by shared underlying genetic variants (Visscher et al., 2017). This phenomenon in which genetic variants affect multiple traits or diseases is called pleiotropy. Several methods have been developed to study pleiotropic SNPs by exploring the genetic relationship of multiple phenotypes. Broadly, these approaches can be categorized into three major groups. The first method is genetic correlation, which aims to quantify the similarity of the genetic effects on pairwise traits using GWAS summary statistics such as LD-score regression (Bulik-Sullivan et al., 2015b) or from individual genotype data with GCTA GREML (Lee et al., 2012). With large population sizes, these methods can accurately partition variance into a shared genetic component but do not reveal the genetic variants driving the genetic correlation. Genome-wide cross-trait analysis (Zhu et al., 2018) has emerged as a means to follow up such results, but these univariate meta-analyses of two traits requires genome wide summary statistics for both traits, can suffer from effect size heterogeneity in combining results from disparate traits, and cannot be easily applied to thousands of traits at once. The second approach is colocalization, which estimates how well the GWAS signals from two signals overlap in a given region while revealing plausibility of individual causal variants (Giambartolomei et al., 2014). These two methods have successfully identified novel genetic connections across distant traits as well as pleiotropic genomic regions but have generally been used independently of each other. Finally, perhaps the most intuitive approach, is quantifying cross-phenotype SNPs that are shared across multiple phenotypes. In its simplest form, a phenome-wide association study takes a single SNP and examines the significance of association across many traits, often from electronic medical record (Denny et al., 2010). Valuable websites, including PhenoScanner (Staley et al., 2016), GRASP (Leslie et al., 2014), and GeneATLAS (Canela-Xandri et al., 2018) have integrated thousands of GWAS studies with billions of SNP-traits associations and allow users to query individual SNPs across the phenome. However, such PheWAS approaches do not leverage shared genetic architecture that extends beyond individual SNPs and do not take advantage of LD information.

Motivated to simultaneously connect human phenotypes with shared genetic architecture and to identify the precise loci driving this similarity, we previously developed a method, CPAG (Cross-phenotype Analysis of GWAS), which estimated phenotype similarity of NHGRI-EBI GWAS catalog traits based on shared genetic associations (Wang et al. 2015). CPAG utilized cross-phenotype SNP associations to cluster traits into groups that were consistent with pre-defined categories and discovered novel pleiotropic SNPs connecting Crohn’s disease and the fatty acid palmitoleic acid. However, CPAG could not scale sufficiently to keep up with the massive increase in the scope and scale of GWAS (facilitated through increasing use of electronic medical record (EMR)-based GWAS of huge cohorts) and the deeper phenotyping of molecular and cellular traits that can provide insight into mechanisms of pathophysiology of disease. Here, we introduce iCPAGdb, a new cross-phenotype analysis platform with improved identification of shared loci using pre-computed ancestry-specific LD databases and a more efficient algorithm for capturing cross-phenotype associations. These improvements facilitated integration of the NHGRI-EBI GWAS catalog with large datasets of plasma and urine metabolites and cellular host-pathogen traits. Such integration of pleiotropic analyses using GWAS datasets that include intermediate traits across biological scales are crucial for moving from lists of associated SNPs to understanding the pathophysiology of complex diseases. Finally, iCPAGdb allows users to upload their own GWAS summary statistics via web interface (http://cpag.oit.duke.edu) to identify and explore shared SNPs between their own GWAS and a deep catalog of 4418 molecular, cellular, and disease phenotypes. Using a GWAS of severe COVID-19 as the querying phenotype in iCPAGdb revealed shared SNPs associated with idiopathic pulmonary fibrosis and plasma protein levels of CD209, a possible receptor for SARS-CoV-2.

## Results

### iCPAGdb: An atlas for discovery of cross-phenotype associations

We created iCPAGdb to facilitate exploration of cross-phenotype associations of human phenotypes and discovery of shared genetics connecting traits that were previously not known to be related. iCPAGdb utilizes 85639 SNP-trait associations (p < 5 x 10^−8^) across 3793 traits from the NHGRI-EBI GWAS catalog, incorporates additional GWAS datasets (see below and Table 1), and allows for uploading and analysis of user GWAS summary statistics (Fig. 1A). In contrast, the original CPAG (published in 2015) used only 14198 SNP-trait associations for 887 traits from the NHGRI-EBI GWAS catalog.

**Table 1.** **A summary of GWAS data in iCPAGdb. GWAS summary statistics were clumped to include only a lead SNP for each trait locus**.

|  | Type | Traits/Diseases # | SNPs (p < 5e-8) | Trait-SNP associations # | Urls |
| --- | --- | --- | --- | --- | --- |
| NHGRI Catalog | Clinical GWAS | 3793 | 63933 | 85639 | https://www.ebi.ac.uk/gwas/ |
| H2P2 | Molecular/cellular GWAS | 79 (44 flow cytometric phenotypes + 35 cytokines) | 17 | 3489 (p<1e-5) | http://h2p2.oit.duke.edu |
| Blood Metabolites | Molecular GWAS | 491 Blood (453 metabolites + 38 xenobiotics) | 1441 | 2024 | http://metabolomics.helmholtz-muenchen.de/gwas/ |
| Urine Metabolites | Molecular GWAS | 55 Urine | 149 | 171 | http://metabolomics.helmholtz-muenchen.de/gwas/ |
| **Sum** |  | 4418 | 65540 | 91323 |  |

**Figure 1.**
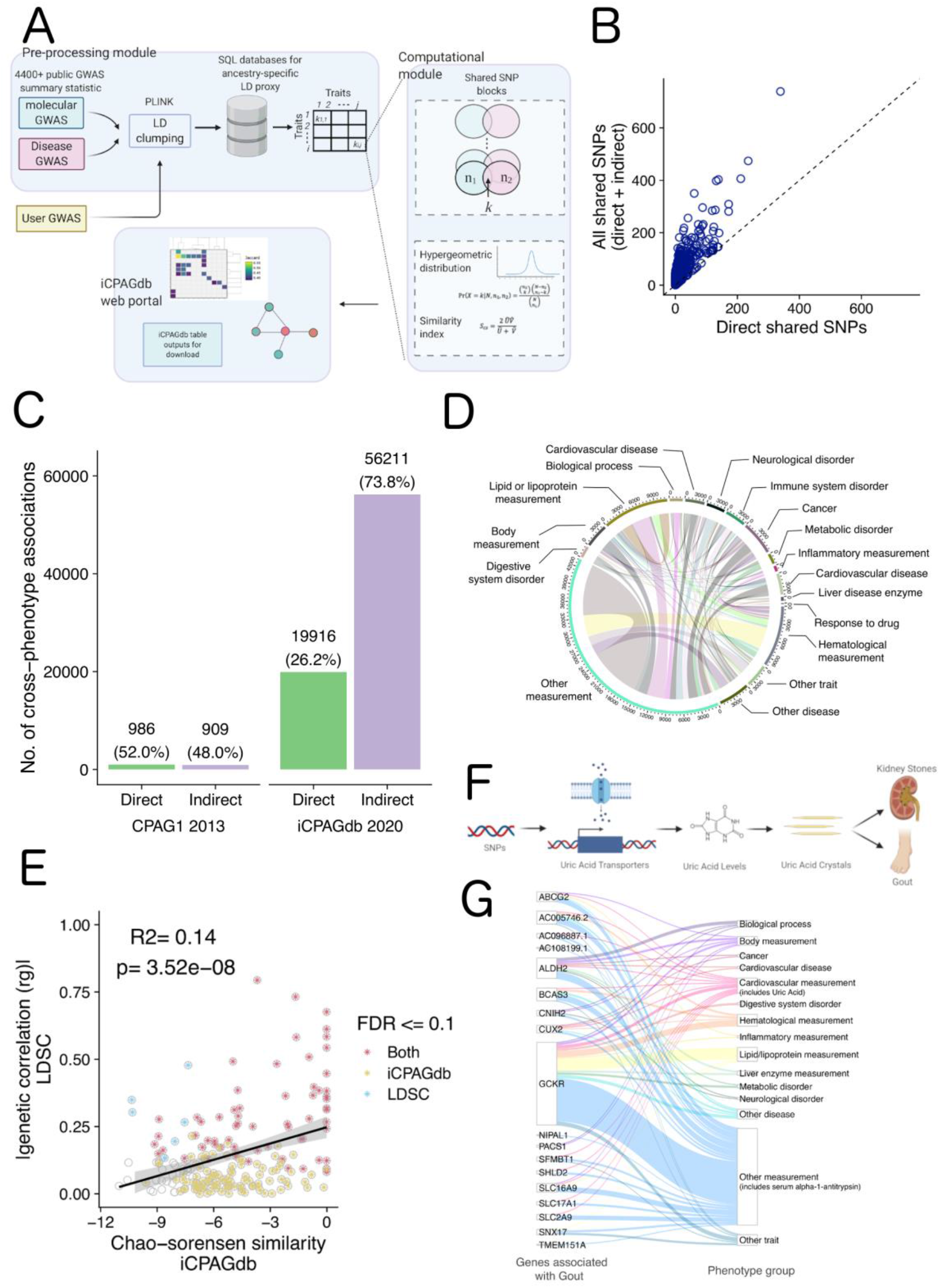
An improved method for finding shared genetic architecture of human traits. **(A)** The overall framework of the iCPAGdb pipeline. GWAS summary statistics (from published GWAS datasets or from user-uploaded GWAS) undergo LD clumping to obtain a lead variant for each signal below a specified p-value threshold. These SNPs are queried against an LD proxy database generated from 1000 Genomes African, Asian, or European population to identify cross-phenotype associations through direct overlap or LD proxy at *R*^2^ > 0.4. Significance of overlap for each trait pair is calculated using Fisher’s exact test. Outputs can be visualized/downloaded from the iCPAGdb web browser. **(B)** Comparison of the number of shared SNPs for each NHGRI-EBI GWAS catalog trait pair identified through direct overlap vs. both direct and indirect (LD-proxy) overlap. **(C)** iCPAGdb detected more significant cross-phenotypes associations than CPAG1 at FDR < 0.1. Expansion of the NHGRI-EBI GWAS catalog and improvements in capturing by LD proxy in iCPAGdb fueled a large increase in detected cross-phenotype associations across human traits. Comparisons between CPAG1 and iCPAGdb on the same 2013 dataset are in Fig. S3. **(D)** Circle plot of cross-phenotype associations detected by iCPAGdb in the NHGRI-EBI GWAS catalog. After excluding compound phenotypes (phenotypes described by NHGRI-EBI GWAS catalog as > 1 comma-separated phenotype in their ontology), a total of 1709 traits involved in a total of 53314 cross-phenotype associations were left. These were categorized into 17 EFO Parental groups. Inner ribbons link phenotypes connected by cross-phenotype associations with the width of ribbon corresponding to the number of cross-phenotype associations. The axis outside the circle represents the cumulative number of associations for each group vs all other groups. **(E)** Comparison of genetic correlation from LD score regression (LDSC) and the Chao-Sorensen similarity index implemented in iCPAG demonstrates significant correlation. The genetic correlation *rg* of 24 diseases/trait were obtained from (Bulik-Sullivan et al., 2015a). Since Chao-Sorensen values are bounded from 0 to 1 and *rg* ranges from -1 to 1, we used the absolute value of *rg* here. Colored * indicates significant trait-pair for LDSC, iCPAGdb, or both at false discovery rate of 0.1. **(F)** A model demonstrating how SNPs regulate uric acid levels to impact the development of kidney stones and gout. **(G)** Riverplot of gout cross-phenotype associations generated from iCPAGdb output shows causal connections, comorbid outcomes, and regulators of disease. Mapped genes for SNPs associated with gout are shown on the left and connected to other NHGRI-EBI GWAS phenotypes grouped by EFO on the right.

Beyond this large expansion in traits and associations, we improved on the original CPAG algorithm by clumping GWAS data from each study (Fig. S1), creating a database of LD values based on 1000 Genomes (Genomes Project et al., 2015), allowing selection of either European, African, or Asian LD structure, and efficiently capturing cross-phenotype associations that are driven by LD proxy (Fig. 1B). For each trait pair, iCPAGdb first selects the lead SNPs from all associated loci at a selected p-value threshold (p < 5 x 10^−8^ was used for analysis of the NHGRI-EBI GWAS catalog; Table S1; Fig. S2). These lead SNPs are compared across the trait pair to count directly shared SNPs. For SNPs that are not directly shared, iCPAGdb then checks an LD database for overlap by LD proxy. For all directly or indirectly shared SNPs, iCPAGdb further forms them into bigger SNP blocks by recursively merging them until each SNP block has no LD proxy with R^2^ >= 0.4 against all others. iCPAGdb improves memory efficiency with built-in functions connecting to SQL GWAS and LD proxy databases and improves computational efficiency and speed by utilizing multiple CPUs. For the NHGRI-EBI GWAS Catalog, the growth of GWAS findings and improvements of iCPAGdb over the previous version of CPAG led to a 27.7-fold increase in direct cross-phenotype associations and a 47.7-fold increase in indirect cross-phenotype associations, many of which would have been missed by the original CPAG algorithm (Fig. 1C, D). Indeed, analyzing the 2013 NHGRI-EBI GWAS catalog with iCPAGdb had little effect on direct associations but increased indirect associations by 76% (Fig. S3).

Results of iCPAGdb are consistent with results from the orthogonal approach of genetic correlation by LD score regression (Bulik-Sullivan et al., 2015b). Comparing the absolute values for genetic correlation of 24 phenotypes from (Bulik-Sullivan et al., 2015a) against a similarity index quantifying the degree of shared SNPs in iCPAGdb revealed that the two are significantly correlated (p=3.52 × 10^−8^; *R*^2^ = 0.14) (Fig. 1E). Nearly all phenotypes (64 of 70) that showed significant correlation by LD score regression also demonstrated a significant excess of shared SNPs in iCPAGdb. The output of iCPAGdb provides the SNPs driving the similarity between the two phenotypes, facilitating follow-up studies. Interestingly, 61% of pairwise comparisons that had significant overlap based on iCPAGdb did not have significant genetic correlation based on LD-score regression. For example, LD-score regression did not detect significant genetic correlation between LDL and HDL cholesterol measurements, but iCPAGdb detected 92 shared SNPs, including 31 by direct overlap where the two phenotypes have the same lead SNPs (p=7.55×10^−195^by Fisher’s exact test; p=1.49×10^−190^ after Benjamini-Hochberg procedure. P-values from iCPAGdb in the remainder of the paper are FDR-corrected for all pairwise comparisons using Benjamini-Hochberg procedure).

### GWAS of varying phenotypic scales reveals shared genetic architecture connecting molecular and cellular traits with human disease

In a previous study (Wang et al., 2015), we defined 4 categories of cross-phenotype associations: 1) SNP similarity between an intermediate trait/risk factor and disease, 2) SNP similarity between a disease and a consequence of disease, 3) SNP similarity between two traits affected by the same gene/pathway, and 4) SNP similarity between two traits affected by the same gene having effects in different tissues or on different pathways. Of these categories, perhaps the most clinically useful is the first category—shared SNPs that connect an intermediate trait to a disease may reveal how molecular or cellular phenotypes mediate some aspect of the pathophysiology of disease. While the NHGRI-EBI GWAS catalog is comprised primarily of case-control GWAS of disease, we detected numerous known shared associations linking a human disease with levels of a metabolite. Metabolites are the substrates, intermediates, and products of cellular metabolism and are routinely already used as biomarkers, such as measuring glucose in diabetes management.

Cross-phenotype associations involving the metabolite uric acid and gout, an inflammatory arthritis driven by excess levels of uric acid (Bodofsky et al., 2020), are illustrative of iCPAGdb’s usefulness. GWAS studies have been conducted on risk of gout (Chen et al., 2018; Lai et al., 2012; Lee et al., 2019; Li et al., 2015; Matsuo et al., 2016; Nakayama et al., 2017; Nakayama et al., 2020; Sulem et al., 2011) as well as uric acid or urate levels (Boocock et al., 2020; Dehghan et al., 2008; Doring et al., 2008; Kamatani et al., 2010; Kottgen et al., 2013; Li et al., 2007; Tin et al., 2019; Tin et al., 2011). Notably, of 31 GWAS loci for gout and 123 GWAS loci for serum uric acid levels at p<5×10^−8^, 13 loci overlap, including 9 loci identified only by LD proxy (nearly 6000-fold enrichment; p=5.9×10^−43^). These loci are spread across 7 chromosomes and include several solute carrier (SLC) and ATP-binding class (ABC) transporters that control urate absorption and secretion. Some of the loci are in close proximity but are counted separately by iCPAGdb, as could occur if different GWAS studies locate nearby peaks that fall below our R^2^>0.4 threshold or if multiple causal signals are located in the same region. These data provide genetic evidence for the well-known causal role of excess uric acid in gout and further reveal multiple genes that may serve as therapeutic targets. Inhibitors of renal uric acid reabsorption through URAT1 (*SLC22A12*) are commonly used in treating gout (Dong et al., 2019), but additional transporters implicated through human genetics may also prove to be useful drug targets. Beyond uric acid levels, GWAS of kidney stones (Howles et al., 2019; Oddsson et al., 2015; Thorleifsson et al., 2009), a second manifestation of elevated uric acid levels, also share associated SNPs with gout (3 shared loci, all identified by proxy on chromosomes 2, 4, and 17; p=5.2×10^−9^). Finally, gout shares 2 loci (out of 5 from (Setoh et al., 2015; Suhre et al., 2017)) with levels of serum alpha-1-antitrypsin, an anti-inflammatory endogenous protease inhibitor (p=9.3×10^−7^), providing a human genetic rationale for the use of alpha-1-antitrypsin-based therapeutics in acute gouty flares (as has been demonstrated to be efficacious in mice (Joosten et al., 2016)). Thus, examining the gout cross-phenotype associations revealed causal connections, comorbid conditions with shared etiology, and factors that modulate inflammation in the disease (Fig. 1F, G).

Shared genetic associations reveal other well-known molecular and cellular disease relationships such as LDL cholesterol levels with cardiovascular disease (1.24×10^−81^) and Alzheimer’s disease (p=4.8×10^−17^) as well as glucose with type II diabetes mellitus (p=1.5×10^−40^). Other cross-phenotype associations highlight genetic variation that can extend our knowledge. For example, cross-phenotype associations were found between malaria (Band et al., 2013; Jallow et al., 2009; Malaria Genomic Epidemiology, 2019; Malaria Genomic Epidemiology et al., 2015; Ravenhall et al., 2018; Timmann et al., 2012) and red blood cell distribution width (Astle et al., 2016; Chen et al., 2020; Chen et al., 2013; Fatumo et al., 2019; Kichaev et al., 2019) (p=1.3×10^−9^). This overlap is driven by well-known genetic variation in the beta-hemoglobin gene (*HBB)* and *ABO* blood type affecting malaria risk but also by genetic variation in *ATP2B4* which encodes a calcium transporter. To the best of our knowledge, whether size of red blood cells impacts susceptibility to malaria parasites has not been examined. These cross-phenotype associations demonstrate the promise of this approach for revealing novel relationships that can be mined through iCPAGdb.

### Expansion of iCPAGdb to additional datasets of molecular and cellular traits

The above examples of clinically relevant cross-phenotype associations involving metabolite and cellular phenotypes motivated expansion of iCPAGdb to additional datasets. We used three datasets to provide molecular and cellular traits to our analysis: 491 metabolites and xenobiotics in blood (Shin et al., 2014) and 55 metabolites in urine (Raffler et al., 2015), both from the Metabolomics GWAS Server (http://metabolomics.helmholtz-muenchen.de/gwas/index.php), and 79 cellular host-pathogen interaction traits from our dataset of cellular host-pathogen interaction GWAS, H2P2 (Wang et al., 2018). iCPAGdb revealed many connections between these molecular/cellular datasets and the NHGRI-EBI GWAS catalog (Fig. 2A; Table S2).

**Figure 2.**
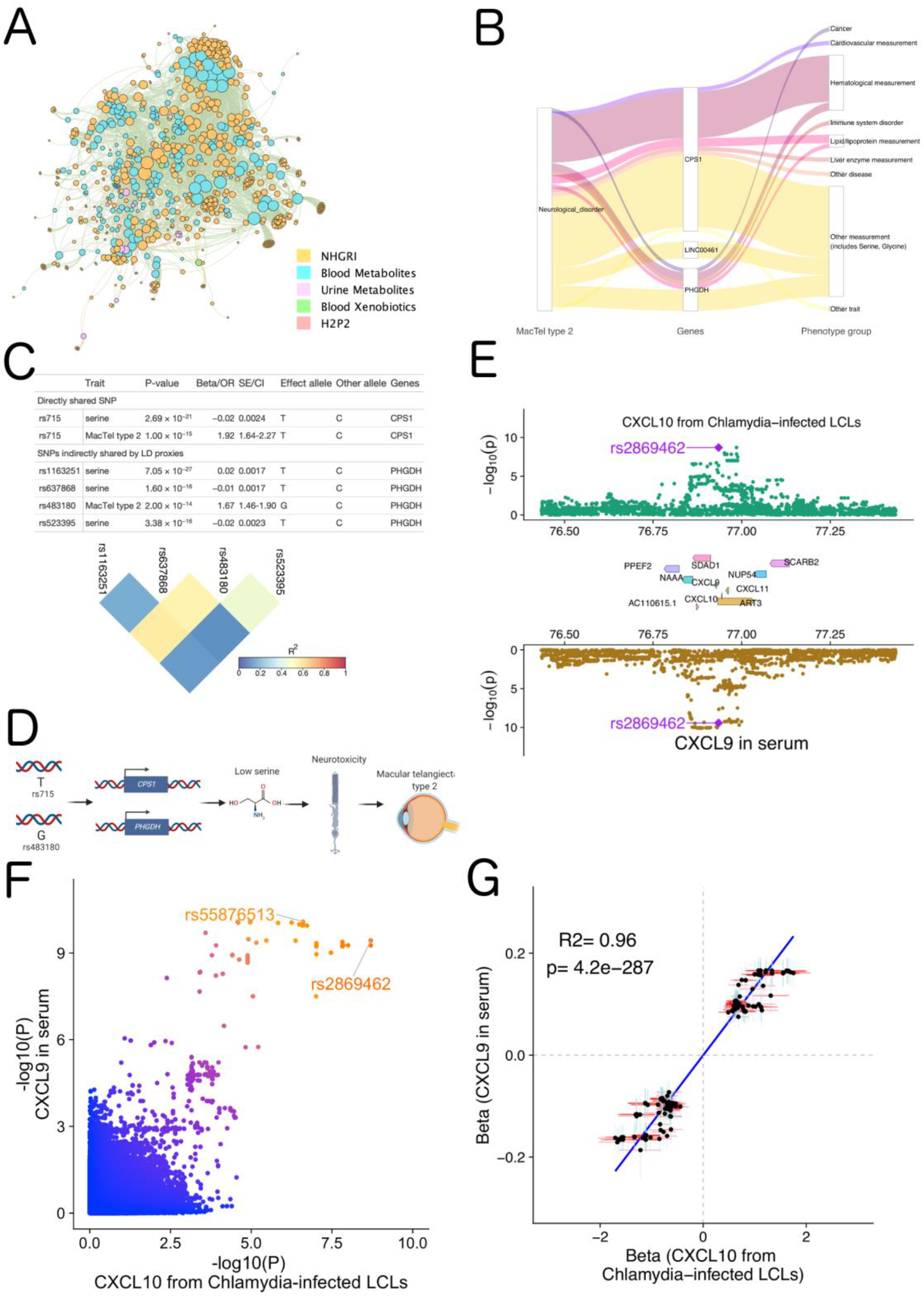
iCPAGdb integrates GWAS of different scales to reveal biological insight. **(A)** Multi-dataset network of cross-phenotype associations detected by iCPAGdb. Phenotypes that demonstrated significant overlap (FDR ≤ 0.1) are color-coded in the indicated colors. **(B)** Riverplot of macular telangiectasia type 2 (MacTel type 2) cross-phenotype associations generated from iCPAGdb shows causal connections, comorbid outcomes, and regulators of disease. **(C)** Cross-phenotype associations connecting MacTel type 2 and serine. One locus demonstrated direct SNP overlap (rs715). A second locus demonstrated indirect overlap based on 4 SNPs in LD as visualized in the heatmap color-coded by LD. **(D)** A model for how SNPs regulate serine levels to impact pathogenesis of MacTel type 2 based on iCPAGdb and prior work described in the text. **(E)** Regional Miami colocalization plot demonstrates a genetic locus that impacts both CXCL10 level in lymphoblastoid cell lines following *Chlamydia trachomatis* infection and CXCL9 (MIG) levels in whole blood. **(F)** Comparison of -log10(p value) for GWAS of *CXCL10* following *Chlamydia trachomatis* infection and levels of *CXCL9* (MIG) in whole blood. The lead SNP in the region for each phenotype is marked. **(G)** Scatter plot demonstrates a highly positive correlation of the effect coefficients of cellular *CXCL10* after *Chlamydia trachomatis* infection and of SNPs associated with blood *CXCL9* levels. Each dot represents a SNP which has p value < 0.01 for both phenotypes. A total of 413 SNPs from a 4-mb window surrounding the leading SNP rs2869462 was selected. The blue vertical or red horizontal bar shows the standard error of the beta value for each SNP.

Cross-phenotype associations with macular telangiectasia (MacTel) type 2, a disease characterized by loss of central vision due to alterations in blood vessels in the macula of the eye, confirmed the importance of the amino acid serine (Fig. 2B). A GWAS of MacTel type 2 uncovered 3 genome-wide significant loci and the authors noted that two of these loci were involved in serine/glycine metabolism, with the alleles associated with low glycine and serine conferring increased risk of MacTel type 2 (Scerri et al., 2017). The authors speculated that the low serine levels could lead to high levels of ammonia and glutamate causing neurotoxicity and stress-induced angiogenesis (Scerri et al., 2017). Gantner et al. have since provided evidence that low serine levels result in elevated levels of deoxysphingolipids to trigger cell death in photoreceptors (Gantner et al., 2019). iCPAGdb rediscovered the connection of two loci being shared between serine in serum (measured by (Shin et al., 2014)) and risk of MacTel (Fig. 2C, D; p=4.0×10^−7^; 99,010-fold enrichment). iCPAGdb also revealed 7 other serum metabolites including glycine that shared an association with rs715 but not with the second MacTel locus. While serine was not part of the urine metabolomics dataset, iCPAGdb did detect overlap of glycine in urine and MacTel type 2 (p=0.01).

We also included host-pathogen traits from H2P2, a cellular GWAS we previously carried out using 528 lymphoblastoid cell lines (LCLs) exposed to 7 different pathogens (Wang et al., 2018). Notably, unlike the metabolomics datasets, H2P2 identified SNPs associated with traits at baseline and in response to stimuli. Further, as pathogens have likely been drivers of human evolution (Fumagalli et al., 2011; Pittman et al., 2016), comparing H2P2 to human disease GWAS may reveal unintended consequences of past pandemics on the human genome. Previously, we reported colocalization of a locus regulating CXCL10 levels following *Chlamydia trachomatis* infection (rs2869462) and risk of inflammatory bowel disease (Wang et al., 2018). iCPAGdb revealed shared genetic variants for this H2P2 phenotype and blood levels of CXCL9 (MIG) (Ahola-Olli et al., 2017) (Fig. 2E; p=0.04). P-values for the two associations are strongly correlated (Fig. 2F), and the effect size for SNPs associated with both chemokines are significantly positive correlated (Fig. 2G). We utilized COLOC, which uses a Bayesian framework to determine whether GWAS signals in the same region are likely due to the same causal variant (Giambartolomei et al., 2014). The posterior probability that both CXCL10 protein levels from cells and CXCL9 levels in blood share the same causal variant is 0.90 (Table 2), with rs2869462 identified as the most likely causal SNP (Table S3). The genes encoding these two chemokines are adjacent to each other on chromosome 4, and this result points to variants regulating expression of both genes that will make it challenging to disentangle their effects in disease.

**Table 2.** COLOC analysis output. PP3 is the posterior probability for the model where the two traits have independent causal variants. PP4 is the posterior probability for the model where the two traits share a single causal variant.

| Trait1 | Trait2 | Locus | SNP # | PP3 | PP4 | PP3+PP4 | PP4/PP3 | Lead causal SNP |
| --- | --- | --- | --- | --- | --- | --- | --- | --- |
| CXCL10 level after *Chlamydia* infection | Blood CXCL9 levels | CXCL10 | 1533 | 0.101 | 0.899 | 1.00 | 8.91 | rs2869462 |
| COVID-19 | Plasma CD209 antigen level | ABO | 56 | 0.0159 | 0.984 | 1.00 | 61.72 | rs505922 |
| COVID-19 | idiopathic pulmonary fibrosis | DPP9 | 1233 | 0.00216 | 0.994 | 0.996 | 459.63 | rs12610495 |

### Application of iCPAGdb to COVID-19 reveals susceptibility due to ABO may occur through regulation of CD209

We applied iCPAGdb to a recently published GWAS of severe COVID-19 with respiratory failure (Ellinghaus et al., 2020). While this study focused on two genome-wide significant associations at the ABO locus and in a cluster of chemokine receptors and other genes on Chromosome 3, we relaxed the p-value threshold for iCPAGdb to 1 × 10^−5^, resulting in 24 suggestive loci after LD clumping. Not surprisingly, iCPAGdb revealed that the genome-wide significant association near the blood type locus *ABO* is in LD with multiple other SNPs in this region associated with other human diseases and traits (Fig. 3A; Table S4). This included the classic association with malaria resistance (Timmann et al., 2012), but also less well known associations with duodenal ulcer (Tanikawa et al., 2012), pancreatic cancer (Amundadottir et al., 2009), and heart failure (Shah et al., 2020). Multiple studies have now reported the association of the *ABO* locus with risk of COVID-19 (Ellinghaus et al., 2020; Zhao et al., 2020). The causal effect on COVID-19 may involve A and B antigens on blood cells, antibodies against A and B antigens, the enzymatic activity of the ABO glycosyltransferase on possibly other glycoproteins, or even other genes in the region. Insight into these possible mechanisms was revealed by iCPAGdb, which identified association of this locus with levels of 8 individual proteins in the NHGRI-EBI GWAS catalog. These proteins, all encoded on different chromosomes than ABO, include IL-6, TNF-α, CD209 (DC-SIGN), Tie-1, mannose-binding protein C, FGF23, and clotting factors (factor VIII and vWF). In each of these cases, the association of the locus to both molecular trait and disease provides a plausible causal chain from SNP to cis-effect on *ABO* to trans effect on a protein to severe COVID-19 disease. For example, association with VWF and Factor VIII may indicate ABO affects COVID-19 through regulation of thrombosis, as patients with severe COVID-19 can have thromboembolic complications as part of a hyper-inflammatory state (Wool and Miller, 2020). In fact, both VWF and factor VIII are targets of glycosylation by ABO (Canis et al., 2018; Matsui et al., 1992; Sodetz et al., 1979) and levels of these proteins are reported to be regulated by ABO (Albanez et al., 2016; Gallinaro et al., 2008; Murray et al., 2020; Shima et al., 1995; Song et al., 2015). Further, regulation of levels of IL-6 and TNF-α suggest possible regulation of inflammation, as “cytokine storm” plays an important role during severe COVID-19 (Mangalmurti and Hunter, 2020). Most interestingly, the *ABO* locus is associated with both COVID-19 and CD209 (p=0.008). A preprint recently confirmed this association across populations, and these authors speculated that ABO may affect CD209 levels to regulate SARS-CoV-2 entry (Katz et al., 2020). Indeed, there has since been evidence from two preprints that CD209 can bind to SARS-CoV-2 and can act as a receptor for entry into immune cells (Amraie et al., 2020; Chen et al., 2021).

**Figure 3.**
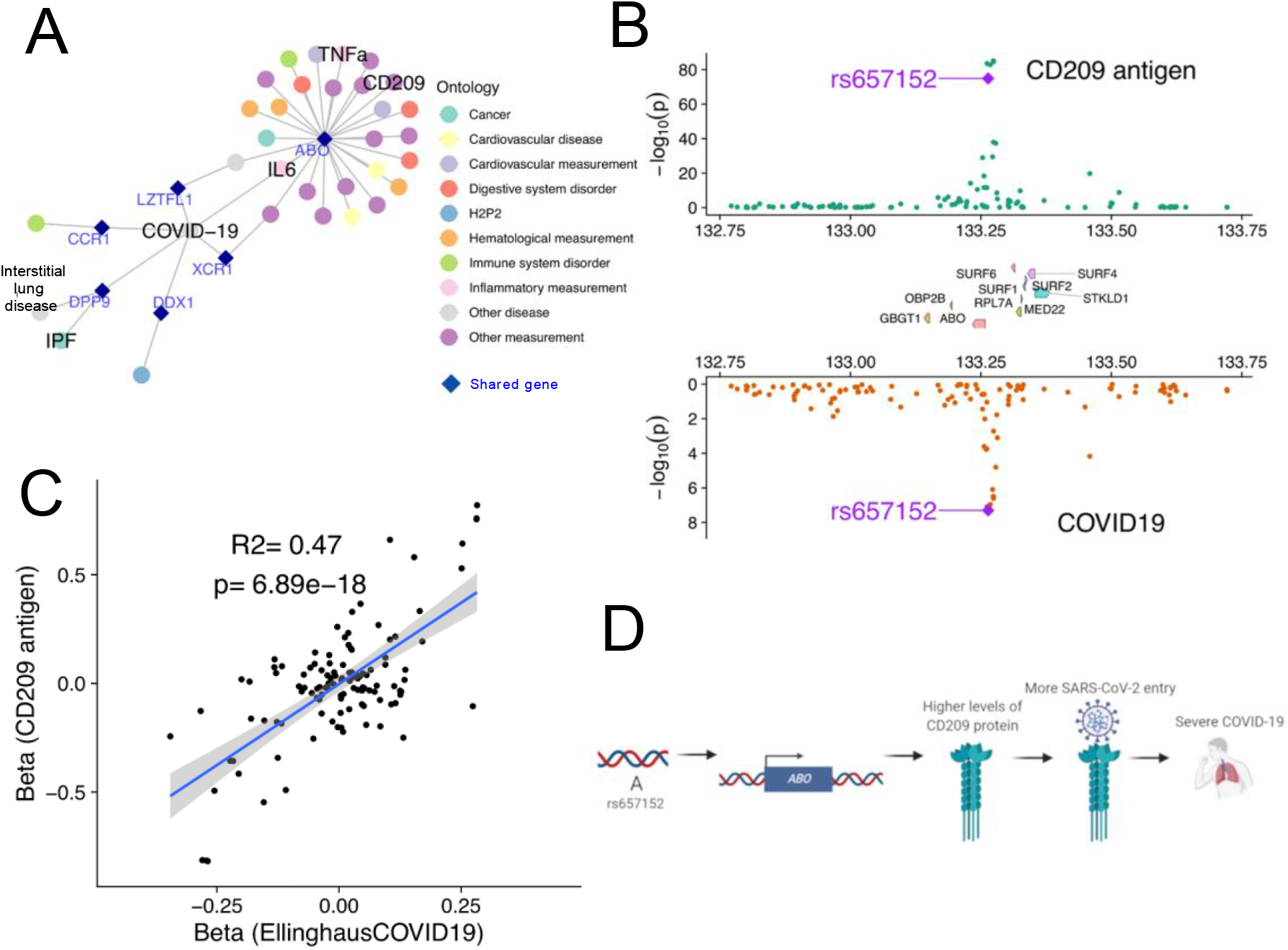
Cross-phenotype association of *ABO* reveals a possible role for CD209 in severe COVID-19. **(A)** A network of genetic associations involving severe COVID-19. Each node represents either a disease/trait (filled circles) or a gene (dark blue diamond). The *ABO* locus was associated with multiple other diseases and levels of specific proteins, while DPP9 connects COVID-19 only with IPF and interstitial lung disease (idiopathic interstitial pneumonia). **(B)** Regional Miami colocalization plot demonstrates the *ABO* locus impacts both CD209 protein levels and risk of severe COVID-19. **(C)** A significant positive correlation for effect size of SNPs in the *ABO* locus on CD209 protein levels and risk of severe COVID-19. **(D)** Model of how *ABO* may affect CD209 and severe COVID-19.

The “A” allele of rs657152 associated with increased risk of COVID-19 with respiratory failure is also associated with increased levels of CD209 (Fig. 3B). We performed colocalization analysis of the GWAS signals for COVID-19 (Ellinghaus et al., 2020) and CD209 protein levels (Suhre et al., 2017). This analysis indicated the two are likely driven by the same causal variants (Fig. 3C; COLOC posterior probability PP4 = 0.98 with the lead causal SNP of rs505922; Table S5). Thus, iCPAGdb and subsequent colocalization analysis support a model where *ABO* regulates CD209 protein levels to impact COVID-19 risk, though much future experimental and clinical studies will be required to fully test this hypothesis (Fig. 3D). The pleiotropic effects of *ABO* on levels of multiple proteins will make defining the mechanism challenging.

### Application of iCPAGdb to COVID-19 reveals a role for DPP9 in regulation of both COVID-19 and idiopathic pulmonary fibrosis

Beyond *ABO*, a locus in the dipeptidyl peptidase 9 (*DPP9*) gene associated at p<1×10^−5^ with severe COVID-19 was identified as being shared with a GWAS of fibrotic idiopathic interstitial pneumonia (Fingerlin et al., 2013) and a recent GWAS of the most severe form of that group of diseases, idiopathic pulmonary fibrosis (IPF) (Allen et al., 2020). rs12610495 was the lead variant for each of these GWAS studies as well as the suggestive peak for severe COVID-19 (p=5.2×10^−6^; (Ellinghaus et al., 2020)). Much evidence has already accumulated that pulmonary fibrosis is a hallmark of severe COVID-19 (Ojo et al., 2020; Shi et al., 2020). While the association of rs12610495 with COVID-19 did not reach genome-wide significance in Ellinghaus et al. 2020, this SNP is in LD with the lead variant from a recent GWAS of critically ill COVID-19 patients that does surpass genome-wide significance (p=3.98×10^−12^; (Pairo-Castineira et al., 2020); R^2^=0.95 in 1000 Genomes European populations). Thus, iCPAGdb alerted us to the importance of a suggestive COVID-19 susceptibility locus that has since been validated in an independent cohort.

We determined that rs12610495 is an eQTL in lung tissue for the gene for *DPP9* (and no other genes in the region) in GTEx (p=4.5×10^−9^; (Bao et al., 2015)), with the “G” allele being associated with lower expression (Fig. 4A). Interestingly, DPP9 is a protease in the same family as DPP4, the receptor for MERS-coronavirus (Raj et al., 2013). Additionally, DPP9 is an inhibitor of inflammasome activation by NLRP3 (Okondo et al., 2017; Okondo et al., 2018; Zhong et al., 2018). Colocalization analysis confirmed the signals from severe COVID-19 and IPF are likely driven by the same causal variant (Fig. 4B; COLOC posterior probability PP4 = 0.994, lead SNP rs12610495; Table S6). Based on these data and the known biology, we developed alternative hypotheses for how this SNP might be regulating risk of severe COVID-19: *DPP9* may be acting as a previously unrecognized receptor for SARS-CoV-2 or it may be inhibiting inflammation during COVID-19 infection. Based on the directionality of effect of rs12610495 on *DPP9* gene expression, the “G” allele should lead to lower *DPP9* expression and less entry if the receptor model is correct. However, the “G” allele is instead associated with increased risk of severe COVID-19 (Fig. 4C). Alternatively, the “G” allele could lead to lower *DPP9* to increase inflammasome activation in lung tissue, a model consistent with “G” increasing risk of severe COVID-19 and this allele also increasing risk of idiopathic pulmonary fibrosis (Fig. 4D).

**Figure 4.**
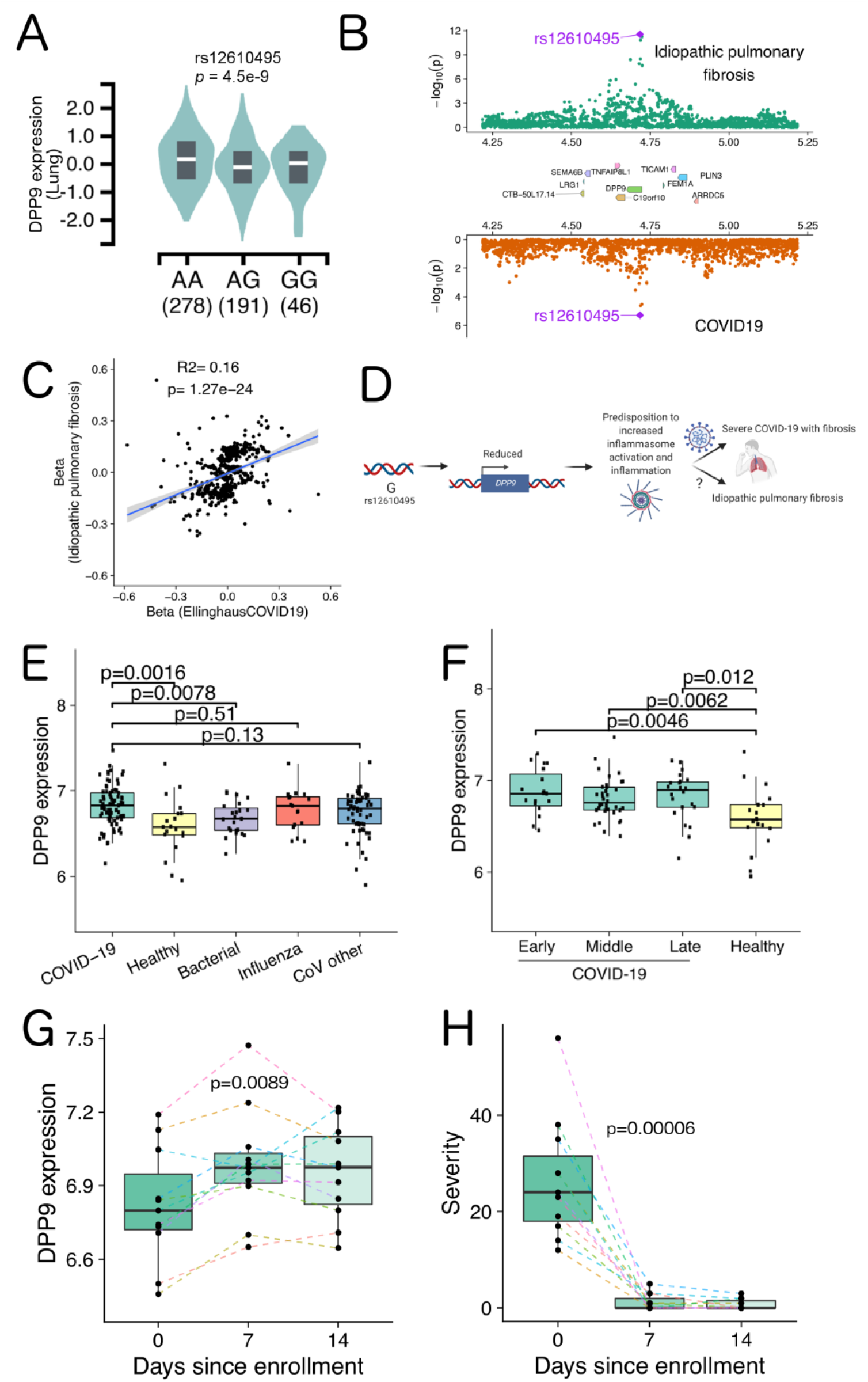
Cross-phenotype analysis and COVID-19 patient transcriptomics reveals a role for *DPP9* in severe COVID-19. **(A)** Lung eQTL data from GTEx shows rs12610495 “G” allele is associated with reduced expression of *DPP9*. **(B)** Regional Miami colocalization plot demonstrates the *DPP9* locus impacts both idiopathic pulmonary fibrosis and risk of severe COVID-19. **(C)** A significant positive correlation for effect size of SNPs in the *DPP9* locus on idiopathic pulmonary fibrosis and risk of severe COVID-19. **(D)** Model of how *DPP9* may affect idiopathic pulmonary fibrosis and risk of severe COVID-19. **(E)** *DPP9* expression in peripheral blood is significantly higher in COVID-19 patients compared to healthy and bacteria-infected patients. The p values were calculated using the Wilcoxon rank-sum test. **(F)** COVID-19 patients demonstrate significantly higher *DPP9* expression compared to healthy controls during early (days 1-10), middle (days 11-20) and late (21+ days) stages of SARS-CoV-2 infection. The p values were calculated using the Wilcoxon rank-sum test. **(G)** *DPP9* demonstrates increased expression during recovery from COVID-19. A total of 11 patients were measured sequentially at enrollment (day 0), day 7, and day 14. The colored dash line connects measurements from the same patient across time points. P value was calculated using Friedman test. **(H)** Decreased symptom severity scores of COVID-19 patients over time. The eleven subjects in G were assessed for symptom severity at day 0, 7 and 14. The colored dash line connects measurements from the same patient across time points. P value was calculated using Friedman test.

To further examine the role of *DPP9* in COVID-19, we analyzed transcriptomics of peripheral blood from COVID-19 patients (McClain et al., 2020). Levels of *DPP9* expression across 46 COVID-19 patients were compared to individuals with seasonal coronavirus, influenza, bacterial pneumonia, and healthy controls. *DPP9* levels were significantly increased in COVID-19 patients compared to the other groups (fold-change = 1.15, p = 0.003 adjusted by Benjamini-Hochberg method). Comparing COVID-19 data vs. each comparator individually revealed that *DPP9* levels were elevated vs. healthy controls (p = 0.0016) and bacterial infection (p = 0.0078) but not influenza or other coronavirus infection (Fig. 4E). This data supports a role for *DPP9* in the host response to viral infections. In examining all samples in the cohort, increased *DPP9* was observed both early and late in COVID-19 infection (Fig. 4F). However, eleven subjects that did not require hospitalization had repeated measurements at day 0 (initial enrollment into the study), day 7, and day 14 that revealed changes in *DPP9* expression as infection resolved. While *DPP9* expression increased from day 0 compared to 7 days and 14 days (Fig. 4G; p = 0.0089), symptom severity dramatically improved over this period (Fig. 4H; p = 0.00006). We speculate that *DPP9* may be induced to effectively turn off the inflammatory response to SARS-CoV-2 to minimize tissue damage and fibrosis. Combined with our human genetic data, these findings suggest that insufficient induction of *DPP9* expression could predispose to severe COVID-19.

### Searching the iCPAGdb web server with user-provided GWAS summary statistics

As the above examples demonstrate, iCPAGdb analysis can rapidly generate hypotheses connecting molecular and cellular traits to human disease. The website allows quick access to the pre-calculated cross-phenotype associations results described in this manuscript. Users can also upload their own GWAS summary statistics for comparing against all 4414 GWAS traits in the iCPAGdb website, facilitating the discovery of new cross-phenotype relationships. Total time for uploading, clumping of summary statistics, and calculation of cross-phenotype associations is typically <2 minutes.

## Discussion

The expansion of GWAS studies to more molecular, cellular, and human disease traits requires the development and implementation of new tools to facilitate drawing meaningful connections between phenotypes and understanding the molecular mechanisms that explain this shared genetic architecture. Our work demonstrates that leveraging available GWAS summary statistics and efficient algorithms of integrating pleiotropic information using ancestry-specific LD structure can rapidly reveal cross-phenotype associations across different phenotypic scales, which can be applied in real-time to better understand ongoing health crises such as the SARS-CoV2 pandemic.

In examining cross-phenotype connections, it is important to carefully examine the overlapping SNPs provided as part of the iCPAGdb output to determine 1) the genome location where the variants are located, as some may be adjacent/overlapping loci in weak LD and not truly distinct, and 2) how well identified GWAS signals from two traits overlap. Indeed, we view iCPAGdb as the first step in a pipeline for gaining greater understanding of any GWAS that then moves to colocalization analysis (see Fig. 2E, 3B, 4B; Table S3, S5, S6), to further dissect GWAS signals in the same region. Making summary statistics more readily available for all GWAS, especially earlier studies in NHGRI-EBI GWAS, would facilitate these validation studies. Finally, functional studies in model systems and clinical studies are needed to test the proposed hypothesis and deeply understand the underlying mechanisms.

While the current web implementation of iCPAGdb uses NHGRI-EBI GWAS catalog (Welter et al., 2014), H2P2 (Wang et al., 2018), and metabolomics GWAS datasets (Raffler et al., 2015; Shin et al., 2014), additional datasets of molecular, cellular, and disease GWAS can be easily added. Analysis of user-uploaded GWAS may be the most useful application of iCPAGdb and will lead to discovery of new connections among human phenotypes to encourage experimental and clinical follow-up studies. Our studies of COVID-19 provide a test case for this and revealed possible mechanisms underlying the associations of severe COVID-19 with *ABO* and *DPP9*.

While our work highlights shared genetic architecture regulating ABO, protein abundance, and COVID-19, much work remains to be done to understand the mechanisms underlying these connections. The ABO locus controls abundance of many proteins. Some of these proteins, such as VWF and Factor VIII, have already been shown to be regulated by glycosylation of ABO (Canis et al., 2018; Matsui et al., 1992; Sodetz et al., 1979). For CD209, *ABO* is a pQTL, but it is unknown whether CD209 protein abundance is regulated by ABO glycosylation. CD209 has a predicted N-linked glycosylation site (N80) and glycosylation has been observed by mass spectrometry (http://glycositeatlas.biomarkercenter.org/glycosites/33001/). Whether human genetic variation also impacts CD209 glycosylation is also an unanswered question. Previous studies have examined protein glycosylation as a GWAS trait, resulting in 16 genome-wide significant loci (Huffman et al., 2011; Lauc et al., 2010; Sharapov et al., 2019), 15 of which have been recently replicated (Sharapov et al., 2020). However, these studies quantified total plasma N-glycans released from proteins and did not specifically quantify glycosylation and glycoforms for individual proteins. Future GWAS quantifying individual glycosylated protein isoforms, as well as other post-translational modifications, may therefore be valuable.

The shared underlying genetic risk factors for IPF and COVID-19 suggest that *DPP9* may have a common role in pathogenesis in these diseases. iCPAGdb was able to identify this connection in the first published COVID19 GWAS despite the *DPP9* allele being below genome-wide significance in that cohort, demonstrating the utility of iCPAGdb in expanding the power of GWAS studies on emerging and understudied diseases. We speculate that characteristics of inflammasome-mediated responses, normally suppressed by high expression of *DPP9*, may predispose to fibrosis. The shared genetic architecture also suggests that therapeutic approaches targeting fibrosis may be beneficial in both conditions. Pirfenidone and Nintedanib are anti-fibrotic FDA-approved drugs used to treat IPF, and our findings support the idea that these drugs may prove beneficial in COVID-19 (Ferrara et al., 2020; George et al., 2020; Seifirad, 2020). As our examination of COVID-19 demonstrates, iCPAGdb is a powerful hypothesis engine that will lead to a deeper understanding of the genetic underpinnings of human disease risk, severity, and drug response.

## Materials and Method

### Collection of GWAS summary statistics

Publicly available GWAS summary statistics were downloaded from the following sources: 3793 traits from NHGRI-EBI GWAS Catalog (version 1.02, downloaded on 2020/08/05), 79 traits from H2P2 cellular GWAS (Wang et al., 2018), 587 traits from human blood circulating metabolites and urine metabolites GWAS (Raffler et al., 2015; Shin et al., 2014). NHGRI-EBI GWAS catalog traits included annotation by Experimental Factor Ontology (EFO). All GWAS data were harmonized to genome coordinates of HG19. In total, we collected 4,225 GWAS traits, and 104,247 Trait-SNPs pairs at a p value threshold of 5 × 10^−8^. A detailed list of trait-SNP pairs at varying p-value threshold can be found in Table 1.

Severe COVID-19 GWAS summary statistics were downloaded from the GRASP website (https://grasp.nhlbi.nih.gov/Covid19GWASResults.aspx) (download date 2020/07/15). Genome coordinates were converted from GRCh38 to HG19 using UCSC liftOver. GWAS summary statistics of IPF were kindly provided by Allen et al. 2020 after requesting access https://github.com/genomicsITER/PFgenetics.

### LD clumping

GWAS summary statistics were individually pre-processed by LD clumping using *PLINK v1*.*9* (Chang et al., 2015), based on genotypes from European populations from the 1000 Genome project. The general PLINK command was “--clump-p1 1e-5 --clump-p2 1 --clump-r2 0.4 --clump-kb 1000”. For NHGRI/EBI GWAS catalog, the index SNPs were selected using the genome-wide significant p value threshold of 5 × 10^−8^ (--clump-p1 5e-8). For molecular and cellular GWAS, we used a varying p-value cutoff from 1 × 10^−3^ to 1 × 10^−5^ for --clump-p1 parameter to choose the index SNPs.

For uploaded GWAS data, iCPAGdb calls on PLINK automatically to perform LD clumping. Users can define the p value for --clump-p1 to select the index SNPs and choose proper LD structure (European, African, or Asian) based on the ancestry of the GWAS.

### LD proxy calculation

To maximize phenotypic associations due to indirect associations, pairwise LD *R*^2^ values were computed for each leading SNP against its surrounding SNPs using the genotypes from the 1000 Genome project (Phase 3 genotypes). Prior to calculation, all SNPs with minor allele frequency less than 0.01 and missingness > 0.1 were removed. *R*^2^ of pairwise SNPs within 10,000 bp windows were then calculated, and only LD proxies with R^2^ > 0.4 were retained in further analysis. The PLINK parameters for calculating LD was “--ld-window-kb 1000 –ld-window 10000 –keep-allele-order –r2 in-phase with-freqs gz”.

Since GWAS may be performed on diverse populations from different ancestry or continents, we calculated ancestry-specific LD proxies for European, African, and Asian populations separately. European population included 503 samples from 5 populations (CEU, TSI, FIN, GBR, IBS), African included 661 samples from 7 populations (YRI, LWK, GWD, MSL, ESN, ASW, ACB), and Asian population included 504 samples from 5 populations (CHB, JPT, CHS, CDX, and KHV). We filtered genotypes for each ancestry population by minor allele frequency more than 0.01 and retained only biallelic SNPs. SNPs which have same genome coordinates were merged using “—merge-equal-pos”. For duplicated SNPs with same variant rsID, we kept only the first variant by using “--rm-dup force-first” using PLINK 2.0,

### Cross-phenotype SNP analysis

Cross-phenotype SNPs were used to quantify the similarity of different traits. Cross-phenotype loci were identified as leading SNPs and/or their LD proxies having statistically significant associations with more than one trait/disease. If two traits shared a common leading SNP, we termed this “direct association”. If a leading SNP was associated with one trait, while its LD proxy SNPs were associated with another trait, we called this “indirect association”. If any shared SNP was in LD with another SNP with R^2^ > 0.4, these SNPs were merged into a SNP block until no further LD was found across shared SNP/LD pairs.

The significant association among each trait pair were using the hypergeometric distribution.

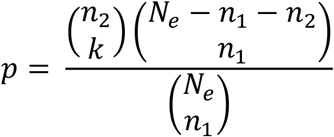

Where *N*_*e*_ is the effective number of independent SNPs in the selected population, the *n*_1_ and *n*_2_ are the number of independent SNPs associated with trait 1 and trait2, and *k* is the number of independent SNPs blocks. The effective number of independent SNPs for European, African and Asian population were obtained from Table 4 from (Li et al., 2012).

The significance of associations for all trait pairs was further corrected for all possible pairwise comparisons using the Benjamini-Hochberg and Bonferroni methods for multiple test correction. A false discovery rate of 0.1 was chosen to identify significantly correlated trait pairs.

### Comparison to LDSC

Bulik-Sullivan et al. (Bulik-Sullivan et al., 2015b) developed an innovative and unbiased method, LDSC, to estimate genetic correlation using GWAS summary statistic for all measured SNPs. Their model calculated the LD scores for a variant against all other variants in a 1 centimorgan window and hypothesized that SNPs with higher LD scores are tagged to a risk-conferring variant, and the genetic correlation among traits can be calculated by normalizing genetic covariance of SNP heritability. With this method, they estimated 276 genetic correlations for 24 diseases/traits based on full GWAS summary statistic (Bulik-Sullivan et al., 2015a). To evaluate the power of iCPAGdb, we calculated the genetic associations on the same 24 GWAS traits. For each trait pair, only SNPs associated with each trait passing genome wide significant threshold (5 x 10^−8^) were used by iCPAGdb. We quantified the strength of cross-phenotype similarity for each trait pair using Chao-Sorensen similarity index. Since the p values from (Bulik-Sullivan et al., 2015a) were not corrected by multiple test correction, we calculated the p values for *r*_*g*_ using R “p.adjust” function with a total number of 276 comparisons.

### Colocalization analysis

To evaluate whether the associations of GWAS trait pairs identified by iCPAGdb were due to sharing the same causal variants, we performed colocalization analysis using R COLOC packages (Giambartolomei et al., 2014). COLOC uses a Bayesian framework to estimate the posterior probability that two GWAS traits share two independent causal signals (PP3) or shares a single casual variant (PP4) in the selected genome region. For each trait pair evaluated by COLOC, SNPs within 200 kb window from the lead SNP were included. Since COLOC requires minor allele frequency (MAF) for each SNP in both GWAS studies, when MAF was not available, we calculated the MAF using European populations from the 1000 Genome Project. We ran COLOC “*coloc*.*abf*” function using the default prior parameters, p1 = 1×10^−4^, p2 = 1×10^−4^, and p12 = 1×10^−5^. We also ran built-in “*sensitivity*” function to evaluate the robustness of predefined priors, and all tests suggested that default prior parameters are robust, therefore, we ran all colocalization analyses with default priors values.

### COVID-19 transcriptomic analysis

As described in (McClain et al., 2020), samples were collected as part of the Molecular and Epidemiological Study of Suspected Infection (MESSI) which was conducted at Duke University Health System (DUHS) and the Durham Veterans Affairs Health Care System (DVAHCS). The study was approved by each institution’s IRB. Informed consent was obtained from all subjects or their legally authorized representatives, and informed consent were collected for all subjects. SARS-CoV-2 RT-PCR testing was used to confirm infection status. A total of 46 subjects were analyzed, 14 of which were assayed at more than 1 timepoint. In total, 77 samples were assayed. Subjects were divided into early (≤10 days), middle (11-21 days), and late (>21 days) stage based on duration of symptoms. Participant self-reported symptoms were recorded at each timepoint for 39 symptom categories. Each symptom was scored on a scale of 0–4, with 0 indicating not present, 1 mild, 2 moderate, 3 severe, and 4 very severe symptoms. Daily symptom severity (sum of symptom scores for all symptoms) was determined for each timepoint. At enrollment (Day 0), date of symptom onset was determined, and an initial symptom survey recorded maximum score for each symptom category between symptom onset and study enrollment. Total RNA was extracted from peripheral whole blood, and cDNA libraries prepared using NuGEN Universal Plus mRNA-seq with AnyDeplete Globin reduction were sequenced on the Illumina NovaSeq 6000, as described (McClain et al., 2020). In brief, STAR v 2.7.1 (Dobin et al., 2013) was used to align the short reads and generate the count matrix. The count matrix was further normalized using TMM method (Robinson and Oshlack, 2010) and log2 transformed. Associations were performed with generalized linear models (LIMMA, (Ritchie et al., 2015)) and corrected for multiple testing using the Benjamini-Hochberg method (Benjamini and Hochberg, 1995). Analysis of *DPP9* was carried out in R, and p values were calculated using the Wilcoxon rank-sum test.

### iCPAGdb software and website implementation

iCPAGdb is comprised of two core parts, the back-end computation and the front-end web browser. The back-end was written in python v3.6 with utilization of SQLite. SQLite tables were constructed for harmonized GWAS datasets and LD tables for different populations and are accessed using python sqlite3 package. The GWAS table stores clumped GWAS summary statistic, including trait name, trait sources, SNPs’ rsID, beta values, standard error/standard deviation of beta, effective allele, and p values. The ancestry-specific LD proxy tables contain pairwise SNPs’ rsID and *R*^2^ values (*R*^2^ >= 0.4) for different populations. All SQLite tables were indexed on unique combinations of SNP and trait or SNP pairs for LD proxy tables, which greatly reduces the searching time. To further increase calculation speed, the core cross-phenotype analysis part of iCPAGdb is parallelized by utilizing multiple threads.

Primary software components for the web portion of iCPAGdb are the R statistical programming language (Team, 2020), the R package Shiny (v1.5.0) for interaction of web pages with R scripts (Cheng et al., 2020), Shiny Server as a 24/7 multi-user platform to make Shiny apps publicly accessible (RStudio, 2020), the database environment SQLite for efficient cient querying of GWAS and CPAG results (Hipp, 2020), and the R package RSQLite to execute SQL queries from within R scripts (Muller et al., 2020). The results of a CPAG execution are read by the R script, processed, and presented to the viewer in various tables and graphs on a web page. The iCPAGdb website is currently loaded with associations across more than 4400 public GWAS datasets that can be browsed and searched in “Review” mode. The user requests an existing CPAG result set from which a corresponding table and heatmap are generated and displayed. Various filtering and graph construction controls are available for iterative sub-setting of data and selection of significance measure and number of top signicant phenotype pairs to plot. The “Download” button enables the researcher to make a local copy of records appearing in the currently displayed results table. Important packages used in this mode are DT for construction of and interaction with tables and ggplot2, plotly, and heatmaply for basic plotting, interactive plotting (hover labels), and heatmap generation, respectively. The web browser also allows users to upload their own GWAS summary data, and iCPAGdb will automatically perform LD clumping based on selected population and generate an atlas of connections for the user’s GWAS against > 4400 GWAS traits in the database. In this “Upload” mode, the user browses filles on a local computer, selects a properly formatted GWAS result file of interest (containing, for a single phenotype, SNP rsIDs and GWAS p-values), specifies format and column configuration, then uploads the file. Next, CPAG computation parameter values, including iCPAGdb GWAS set to be crossed with, significance thresholds for filtering, and linkage disequilibrium (LD) population are specified. When “Compute CPAG” is pressed, the R script composes a system level command to execute the CPAG (Python) function. The future() function of the R future package (Bengtsson, 2020) combined with a delaying pipe from the promises package execute CPAG operations asynchronously, waiting on completion before resuming R script execution. Typical run time for a single uploaded GWAS that is already clumped to lead variants is <30 seconds. For GWAS summary statistics including all SNPs in a study, run time is typically < 2 minutes. The results are available with downloadable tables and figures. Additional information on webapp is in Supplemental Note.

Web resources:

iCPAGdb: http://cpag.oit.duke.edu

NHGRI GWAS Catalog: https://www.ebi.ac.uk/gwas/

H2P2 cellular GWAS: http://h2p2.oit.duke.edu

Human metabolite GWAS summary statistics: http://metabolomics.helmholtz-muenchen.de/gwas/index.php?task=download

COVID-19 GWAS summary statistics from Ellinghaus et al. (2020): https://grasp.nhlbi.nih.gov/Covid19GWASResults.aspx

IPF GWAS: download link was obtained by applying for access following the collaborative protocol from https://github.com/genomicsITER/PFgenetics

Tools for visualization:

R packages:

ggplot2: https://cran.r-project.org/web/packages/ggplot2/

gggene: https://cran.r-project.org/web/packages/gggenes/index.html

tidygraph: https://cran.r-project.org/web/packages/tidygraph/

ggnetwork: https://cran.r-project.org/web/packages/ggnetwork/

circlize: https://cran.r-project.org/web/packages/circlize/

ggpubr: https://cran.r-project.org/web/packages/ggpubr/

DT: https://cran.r-project.org/web/packages/DT

plotly: https://cran.r-project.org/web/packages/plotly/

heatmaply: https://cran.r-project.org/web/packages/heatmaply/

promises: https://CRAN.R-project.org/package=promises

Further information and requests for resources should be directed to and will be fulfilled by the Lead Contact, Dennis C. Ko. All iCPAGdb output described in this manuscript are available for browsing from http://cpag.oit.duke.edu. Supplemental files also contain iCPAGdb output and COLOC analysis results. Code is available at GitHub https://github.com/tbalmat/iCPAGdb.

## Supporting information

TableS1_iCPAGdb_NHGRI

TableS2_iCPAGdb_molecularvsNHGRI

TableS3_coloc_CXCL10vsCXCL9

TableS4_iCPAGdb_COVID19

TableS5_coloc_ABO

TableS6_coloc_DPP9

SupplementalNote_iCPAGdbWebApp

## Data Availability

Requests for resources should be directed to and will be fulfilled by the Lead Contact, Dennis C. Ko. All iCPAGdb output described in this manuscript are available for browsing from http://cpag.oit.duke.edu. Supplemental files also contain iCPAGdb output and COLOC analysis results. Code is available at GitHub https://github.com/tbalmat/iCPAGdb.

https://github.com/tbalmat/iCPAGdb

## Acknowledgements

LW, TJB, AI, MRD, ERH, and DCK were supported by NIH R21AI133305. LW, ALA, and DCK were supported by NIH R01AI118903. FJC, RH, TWB, MTM, XS, ELT, ERK, and CWW were supported by DARPA/USMRAA W911NF1920111. We thank Benjamin Schott, Jeffrey Bourgeois, and Kyle Gibbs for thoughtful discussions. We thank Dr. Richard J. Allen and colleagues for sharing idiopathic pulmonary fibrosis GWAS summary statistics.

## Author Contributions

LW and DCK conceived of the study. LW, TJB, ERH, AI, MRD, ERH, and DCK developed iCPAGdb. LW, TJB, FJC and RH carried out computational analysis. LW, ALA, and DCK analyzed iCPAGdb results. MTM, FJC, RH, TWB, XS, GSG, ELT, ERK, and CWW carried out the COVID-19 transcriptomics study and helped design subsequent analysis carried out by LW. All authors contributed to the manuscript.

## References

Ahola-Olli, A.V., Wurtz, P., Havulinna, A.S., Aalto, K., Pitkanen, N., Lehtimaki, T., Kahonen, M., Lyytikainen, L.P., Raitoharju, E., Seppala, I., et al. (2017). Genome-wide Association Study Identifies 27 Loci Influencing Concentrations of Circulating Cytokines and Growth Factors. Am J Hum Genet 100, 40–50.

Albanez, S., Ogiwara, K., Michels, A., Hopman, W., Grabell, J., James, P., and Lillicrap, D. (2016). Aging and ABO blood type influence von Willebrand factor and factor VIII levels through interrelated mechanisms. J Thromb Haemost 14, 953–963.

Allen, R.J., Guillen-Guio, B., Oldham, J.M., Ma, S.F., Dressen, A., Paynton, M.L., Kraven, L.M., Obeidat, M., Li, X., Ng, M., et al. (2020). Genome-Wide Association Study of Susceptibility to Idiopathic Pulmonary Fibrosis. Am J Respir Crit Care Med 201, 564–574.

Amraie, R., Napoleon, M.A., Yin, W., Berrigan, J., Suder, E., Zhao, G., Olejnik, J., Gummuluru, S., Muhlberger, E., Chitalia, V., et al. (2020). CD209L/L-SIGN and CD209/DC-SIGN act as receptors for SARS-CoV-2 and are differentially expressed in lung and kidney epithelial and endothelial cells. bioRxiv.

Amundadottir, L., Kraft, P., Stolzenberg-Solomon, R.Z., Fuchs, C.S., Petersen, G.M., Arslan, A.A., Bueno-de-Mesquita, H.B., Gross, M., Helzlsouer, K., Jacobs, E.J., et al. (2009). Genome-wide association study identifies variants in the ABO locus associated with susceptibility to pancreatic cancer. Nat Genet 41, 986–990.

Astle, W.J., Elding, H., Jiang, T., Allen, D., Ruklisa, D., Mann, A.L., Mead, D., Bouman, H., Riveros-Mckay, F., Kostadima, M.A., et al. (2016). The Allelic Landscape of Human Blood Cell Trait Variation and Links to Common Complex Disease. Cell 167, 1415–1429 e1419.

Band, G., Le, Q.S., Jostins, L., Pirinen, M., Kivinen, K., Jallow, M., Sisay-Joof, F., Bojang, K., Pinder, M., Sirugo, G., et al. (2013). Imputation-based meta-analysis of severe malaria in three African populations. PLoS Genet 9, e1003509.

Bao, Y., Xu, S., Jing, X., Meng, L., and Qin, Z. (2015). De novo assembly and characterization of Oryza officinalis leaf transcriptome by using RNA-seq. Biomed Res Int 2015, 982065.

Bengtsson, H. (2020). A unifying framework for parallel and distributed processing in r using futures.

Benjamini, Y., and Hochberg, Y. (1995). Controlling the false discovery rate: a practical and powerful approach to multiple testing. J R Stat Soc Ser B Methodol 57.

Bodofsky, S., Merriman, T.R., Thomas, T.J., and Schlesinger, N. (2020). Advances in our understanding of gout as an auto-inflammatory disease. Semin Arthritis Rheum 50, 1089–1100.

Boocock, J., Leask, M., Okada, Y., Asian Genetic Epidemiology Network, C., Matsuo, H., Kawamura, Y., Shi, Y., Li, C., Mount, D.B., Mandal, A.K., et al. (2020). Genomic dissection of 43 serum urate-associated loci provides multiple insights into molecular mechanisms of urate control. Human molecular genetics 29, 923–943.

Bulik-Sullivan, B., Finucane, H.K., Anttila, V., Gusev, A., Day, F.R., Loh, P.R., ReproGen, C., Psychiatric Genomics, C., Genetic Consortium for Anorexia Nervosa of the Wellcome Trust Case Control, C., Duncan, L., et al. (2015a). An atlas of genetic correlations across human diseases and traits. Nat Genet 47, 1236–1241.

Bulik-Sullivan, B.K., Loh, P.R., Finucane, H.K., Ripke, S., Yang, J., Schizophrenia Working Group of the Psychiatric Genomics, C., Patterson, N., Daly, M.J., Price, A.L., and Neale, B.M. (2015b). LD Score regression distinguishes confounding from polygenicity in genome-wide association studies. Nat Genet 47, 291–295.

Bycroft, C., Freeman, C., Petkova, D., Band, G., Elliott, L.T., Sharp, K., Motyer, A., Vukcevic, D., Delaneau, O., O’Connell, J., et al. (2018). The UK Biobank resource with deep phenotyping and genomic data. Nature 562, 203–209.

Canela-Xandri, O., Rawlik, K., and Tenesa, A. (2018). An atlas of genetic associations in UK Biobank. Nat Genet 50, 1593–1599.

Canis, K., Anzengruber, J., Garenaux, E., Feichtinger, M., Benamara, K., Scheiflinger, F., Savoy, L.A., Reipert, B.M., and Malisauskas, M. (2018). In-depth comparison of N-glycosylation of human plasma-derived factor VIII and different recombinant products: from structure to clinical implications. J Thromb Haemost.

Chang, C.C., Chow, C.C., Tellier, L.C., Vattikuti, S., Purcell, S.M., and Lee, J.J. (2015). Second-generation PLINK: rising to the challenge of larger and richer datasets. GigaScience 4, 7.

Chen, C.J., Tseng, C.C., Yen, J.H., Chang, J.G., Chou, W.C., Chu, H.W., Chang, S.J., and Liao, W.T. (2018). ABCG2 contributes to the development of gout and hyperuricemia in a genome-wide association study. Sci Rep 8, 3137.

Chen, M.H., Raffield, L.M., Mousas, A., Sakaue, S., Huffman, J.E., Moscati, A., Trivedi, B., Jiang, T., Akbari, P., Vuckovic, D., et al. (2020). Trans-ethnic and Ancestry-Specific Blood-Cell Genetics in 746,667 Individuals from 5 Global Populations. Cell 182, 1198–1213 e1114.

Chen, Q., Gao, R., and Jia, L. (2021). Enhancement of the peroxidase-like activity of aptamers modified gold nanoclusters by bacteria for colorimetric detection of Salmonella typhimurium. Talanta 221, 121476.

Chen, Z., Tang, H., Qayyum, R., Schick, U.M., Nalls, M.A., Handsaker, R., Li, J., Lu, Y., Yanek, L.R., Keating, B., et al. (2013). Genome-wide association analysis of red blood cell traits in African Americans: the COGENT Network. Human molecular genetics 22, 2529–2538.

Cheng, W., Cheng, J., Allaire, J.J., Xie, Y., and McPherson, J. (2020). Shiny: Web Application Framework for R.

Dehghan, A., Kottgen, A., Yang, Q., Hwang, S.J., Kao, W.L., Rivadeneira, F., Boerwinkle, E., Levy, D., Hofman, A., Astor, B.C., et al. (2008). Association of three genetic loci with uric acid concentration and risk of gout: a genome-wide association study. Lancet 372, 1953–1961.

Denny, J.C., Ritchie, M.D., Basford, M.A., Pulley, J.M., Bastarache, L., Brown-Gentry, K., Wang, D., Masys, D.R., Roden, D.M., and Crawford, D.C. (2010). PheWAS: demonstrating the feasibility of a phenome-wide scan to discover gene-disease associations. Bioinformatics 26, 1205–1210.

Dobin, A., Davis, C.A., Schlesinger, F., Drenkow, J., Zaleski, C., Jha, S., Batut, P., Chaisson, M., and Gingeras, T.R. (2013). STAR: ultrafast universal RNA-seq aligner. Bioinformatics 29, 15–21.

Dong, Y., Zhao, T., Ai, W., Zalloum, W.A., Kang, D., Wu, T., Liu, X., and Zhan, P. (2019). Novel urate transporter 1 (URAT1) inhibitors: a review of recent patent literature (2016-2019). Expert Opin Ther Pat 29, 871–879.

Doring, A., Gieger, C., Mehta, D., Gohlke, H., Prokisch, H., Coassin, S., Fischer, G., Henke, K., Klopp, N., Kronenberg, F., et al. (2008). SLC2A9 influences uric acid concentrations with pronounced sex-specific effects. Nat Genet 40, 430–436.

Ellinghaus, D., Degenhardt, F., Bujanda, L., Buti, M., Albillos, A., Invernizzi, P., Fernandez, J., Prati, D., Baselli, G., Asselta, R., et al. (2020). Genomewide Association Study of Severe Covid-19 with Respiratory Failure. N Engl J Med.

Fatumo, S., Carstensen, T., Nashiru, O., Gurdasani, D., Sandhu, M., and Kaleebu, P. (2019). Complimentary Methods for Multivariate Genome-Wide Association Study Identify New Susceptibility Genes for Blood Cell Traits. Front Genet 10, 334.

Ferrara, F., Granata, G., Pelliccia, C., La Porta, R., and Vitiello, A. (2020). The added value of pirfenidone to fight inflammation and fibrotic state induced by SARS-CoV-2 : Anti-inflammatory and anti-fibrotic therapy could solve the lung complications of the infection? Eur J Clin Pharmacol 76, 1615–1618.

Fingerlin, T.E., Murphy, E., Zhang, W., Peljto, A.L., Brown, K.K., Steele, M.P., Loyd, J.E., Cosgrove, G.P., Lynch, D., Groshong, S., et al. (2013). Genome-wide association study identifies multiple susceptibility loci for pulmonary fibrosis. Nat Genet 45, 613–620.

Fumagalli, M., Sironi, M., Pozzoli, U., Ferrer-Admetlla, A., Pattini, L., and Nielsen, R. (2011). Signatures of environmental genetic adaptation pinpoint pathogens as the main selective pressure through human evolution. PLoS Genet 7, e1002355.

Gallinaro, L., Cattini, M.G., Sztukowska, M., Padrini, R., Sartorello, F., Pontara, E., Bertomoro, A., Daidone, V., Pagnan, A., and Casonato, A. (2008). A shorter von Willebrand factor survival in O blood group subjects explains how ABO determinants influence plasma von Willebrand factor. Blood 111, 3540–3545.

Gantner, M.L., Eade, K., Wallace, M., Handzlik, M.K., Fallon, R., Trombley, J., Bonelli, R., Giles, S., Harkins-Perry, S., Heeren, T.F.C., et al. (2019). Serine and Lipid Metabolism in Macular Disease and Peripheral Neuropathy. N Engl J Med 381, 1422–1433.

Genomes Project, C., Auton, A., Brooks, L.D., Durbin, R.M., Garrison, E.P., Kang, H.M., Korbel, J.O., Marchini, J.L., McCarthy, S., McVean, G.A., et al. (2015). A global reference for human genetic variation. Nature 526, 68–74.

George, P.M., Wells, A.U., and Jenkins, R.G. (2020). Pulmonary fibrosis and COVID-19: the potential role for antifibrotic therapy. Lancet Respir Med 8, 807–815.

Giambartolomei, C., Vukcevic, D., Schadt, E.E., Franke, L., Hingorani, A.D., Wallace, C., and Plagnol, V. (2014). Bayesian test for colocalisation between pairs of genetic association studies using summary statistics. PLoS Genet 10, e1004383.

Hipp, R.D. (2020). SQLite.

Howles, S.A., Wiberg, A., Goldsworthy, M., Bayliss, A.L., Gluck, A.K., Ng, M., Grout, E., Tanikawa, C., Kamatani, Y., Terao, C., et al. (2019). Genetic variants of calcium and vitamin D metabolism in kidney stone disease. Nat Commun 10, 5175.

Huffman, J.E., Knezevic, A., Vitart, V., Kattla, J., Adamczyk, B., Novokmet, M., Igl, W., Pucic, M., Zgaga, L., Johannson, A., et al. (2011). Polymorphisms in B3GAT1, SLC9A9 and MGAT5 are associated with variation within the human plasma N-glycome of 3533 European adults. Human molecular genetics 20, 5000–5011.

Jallow, M., Teo, Y.Y., Small, K.S., Rockett, K.A., Deloukas, P., Clark, T.G., Kivinen, K., Bojang, K.A., Conway, D.J., Pinder, M., et al. (2009). Genome-wide and fine-resolution association analysis of malaria in West Africa. Nat Genet 41, 657–665.

Joosten, L.A., Crisan, T.O., Azam, T., Cleophas, M.C., Koenders, M.I., van de Veerdonk, F.L., Netea, M.G., Kim, S., and Dinarello, C.A. (2016). Alpha-1-anti-trypsin-Fc fusion protein ameliorates gouty arthritis by reducing release and extracellular processing of IL-1beta and by the induction of endogenous IL-1Ra. Annals of the rheumatic diseases 75, 1219–1227.

Kamatani, Y., Matsuda, K., Okada, Y., Kubo, M., Hosono, N., Daigo, Y., Nakamura, Y., and Kamatani, N. (2010). Genome-wide association study of hematological and biochemical traits in a Japanese population. Nat Genet 42, 210–215.

Katz, D.H., Tahir, U.A., Ngo, D., Benson, M.D., Bick, A.G., Pampana, A., Gao, Y., Keyes, M.J., Correa, A., Sinha, S., et al. (2020). Proteomic Profiling in Biracial Cohorts Implicates DC-SIGN as a Mediator of Genetic Risk in COVID-19. medRxiv.

Kichaev, G., Bhatia, G., Loh, P.R., Gazal, S., Burch, K., Freund, M.K., Schoech, A., Pasaniuc, B., and Price, A.L. (2019). Leveraging Polygenic Functional Enrichment to Improve GWAS Power. Am J Hum Genet 104, 65–75.

Kottgen, A., Albrecht, E., Teumer, A., Vitart, V., Krumsiek, J., Hundertmark, C., Pistis, G., Ruggiero, D., O’Seaghdha, C.M., Haller, T., et al. (2013). Genome-wide association analyses identify 18 new loci associated with serum urate concentrations. Nat Genet 45, 145–154.

Lai, H.M., Chen, C.J., Su, B.Y., Chen, Y.C., Yu, S.F., Yen, J.H., Hsieh, M.C., Cheng, T.T., and Chang, S.J. (2012). Gout and type 2 diabetes have a mutual inter-dependent effect on genetic risk factors and higher incidences. Rheumatology (Oxford) 51, 715–720.

Lauc, G., Essafi, A., Huffman, J.E., Hayward, C., Knezevic, A., Kattla, J.J., Polasek, O., Gornik, O., Vitart, V., Abrahams, J.L., et al. (2010). Genomics meets glycomics-the first GWAS study of human N-Glycome identifies HNF1alpha as a master regulator of plasma protein fucosylation. PLoS Genet 6, e1001256.

Lee, M.G., Hsu, T.C., Chen, S.C., Lee, Y.C., Kuo, P.H., Yang, J.H., Chang, H.H., and Lee, C.C. (2019). Integrative Genome-Wide Association Studies of eQTL and GWAS Data for Gout Disease Susceptibility. Sci Rep 9, 4981.

Lee, S.H., Yang, J., Goddard, M.E., Visscher, P.M., and Wray, N.R. (2012). Estimation of pleiotropy between complex diseases using single-nucleotide polymorphism-derived genomic relationships and restricted maximum likelihood. Bioinformatics 28, 2540–2542.

Leslie, R., O’Donnell, C.J., and Johnson, A.D. (2014). GRASP: analysis of genotype-phenotype results from 1390 genome-wide association studies and corresponding open access database. Bioinformatics 30, i185–194.

Li, C., Li, Z., Liu, S., Wang, C., Han, L., Cui, L., Zhou, J., Zou, H., Liu, Z., Chen, J., et al. (2015). Genome-wide association analysis identifies three new risk loci for gout arthritis in Han Chinese. Nat Commun 6, 7041.

Li, M.X., Yeung, J.M., Cherny, S.S., and Sham, P.C. (2012). Evaluating the effective numbers of independent tests and significant p-value thresholds in commercial genotyping arrays and public imputation reference datasets. Human genetics 131, 747–756.

Li, S., Sanna, S., Maschio, A., Busonero, F., Usala, G., Mulas, A., Lai, S., Dei, M., Orru, M., Albai, G., et al. (2007). The GLUT9 gene is associated with serum uric acid levels in Sardinia and Chianti cohorts. PLoS Genet 3, e194.

Malaria Genomic Epidemiology, N. (2019). Insights into malaria susceptibility using genome-wide data on 17,000 individuals from Africa, Asia and Oceania. Nat Commun 10, 5732.

Malaria Genomic Epidemiology, N., Band, G., Rockett, K.A., Spencer, C.C., and Kwiatkowski, D.P. (2015). A novel locus of resistance to severe malaria in a region of ancient balancing selection. Nature 526, 253–257.

Mangalmurti, N., and Hunter, C.A. (2020). Cytokine Storms: Understanding COVID-19. Immunity 53, 19–25.

Matsui, T., Titani, K., and Mizuochi, T. (1992). Structures of the asparagine-linked oligosaccharide chains of human von Willebrand factor. Occurrence of blood group A, B, and H(O) structures. J Biol Chem 267, 8723–8731.

Matsuo, H., Yamamoto, K., Nakaoka, H., Nakayama, A., Sakiyama, M., Chiba, T., Takahashi, A., Nakamura, T., Nakashima, H., Takada, Y., et al. (2016). Genome-wide association study of clinically defined gout identifies multiple risk loci and its association with clinical subtypes. Annals of the rheumatic diseases 75, 652–659.

McCarty, C.A., Chisholm, R.L., Chute, C.G., Kullo, I.J., Jarvik, G.P., Larson, E.B., Li, R., Masys, D.R., Ritchie, M.D., Roden, D.M., et al. (2011). The eMERGE Network: a consortium of biorepositories linked to electronic medical records data for conducting genomic studies. BMC Med Genomics 4, 13.

McClain, M.T., Constantine, F.J., Henao, R., Liu, Y., Tsalik, E.L., Burke, T.W., Steinbrink, J.M., Petzold, E., Nicholson, B.P., Rolfe, R., et al. (2020). Dysregulated transcriptional responses to SARS-CoV-2 in the periphery support novel diagnostic approaches. medRxiv.

Muller, K., Wickham, H., James, D.A., and Falcon, S. (2020). RSQLite: ‘SQLite’ Interface for R.

Murray, G.P., Post, S.R., and Post, G.R. (2020). ABO blood group is a determinant of von Willebrand factor protein levels in human pulmonary endothelial cells. J Clin Pathol 73, 347–349.

Nakayama, A., Nakaoka, H., Yamamoto, K., Sakiyama, M., Shaukat, A., Toyoda, Y., Okada, Y., Kamatani, Y., Nakamura, T., Takada, T., et al. (2017). GWAS of clinically defined gout and subtypes identifies multiple susceptibility loci that include urate transporter genes. Annals of the rheumatic diseases 76, 869–877.

Nakayama, A., Nakatochi, M., Kawamura, Y., Yamamoto, K., Nakaoka, H., Shimizu, S., Higashino, T., Koyama, T., Hishida, A., Kuriki, K., et al. (2020). Subtype-specific gout susceptibility loci and enrichment of selection pressure on ABCG2 and ALDH2 identified by subtype genome-wide meta-analyses of clinically defined gout patients. Annals of the rheumatic diseases 79, 657–665.

Oddsson, A., Sulem, P., Helgason, H., Edvardsson, V.O., Thorleifsson, G., Sveinbjornsson, G., Haraldsdottir, E., Eyjolfsson, G.I., Sigurdardottir, O., Olafsson, I., et al. (2015). Common and rare variants associated with kidney stones and biochemical traits. Nat Commun 6, 7975.

Ojo, A.S., Balogun, S.A., Williams, O.T., and Ojo, O.S. (2020). Pulmonary Fibrosis in COVID-19 Survivors: Predictive Factors and Risk Reduction Strategies. Pulm Med 2020, 6175964.

Okondo, M.C., Johnson, D.C., Sridharan, R., Go, E.B., Chui, A.J., Wang, M.S., Poplawski, S.E., Wu, W., Liu, Y., Lai, J.H., et al. (2017). DPP8 and DPP9 inhibition induces pro-caspase-1-dependent monocyte and macrophage pyroptosis. Nat Chem Biol 13, 46–53.

Okondo, M.C., Rao, S.D., Taabazuing, C.Y., Chui, A.J., Poplawski, S.E., Johnson, D.C., and Bachovchin, D.A. (2018). Inhibition of Dpp8/9 Activates the Nlrp1b Inflammasome. Cell Chem Biol 25, 262–267 e265.

Pairo-Castineira, E., Clohisey, S., Klaric, L., Bretherick, A.D., Rawlik, K., Pasko, D., Walker, S., Parkinson, N., Fourman, M.H., Russell, C.D., et al. (2020). Genetic mechanisms of critical illness in Covid-19. Nature.

Pittman, K.J., Glover, L.C., Wang, L., and Ko, D.C. (2016). The Legacy of Past Pandemics: Common Human Mutations That Protect against Infectious Disease. PLoS Pathog 12, e1005680.

Raffler, J., Friedrich, N., Arnold, M., Kacprowski, T., Rueedi, R., Altmaier, E., Bergmann, S., Budde, K., Gieger, C., Homuth, G., et al. (2015). Genome-Wide Association Study with Targeted and Non-targeted NMR Metabolomics Identifies 15 Novel Loci of Urinary Human Metabolic Individuality. PLoS Genet 11, e1005487.

Raj, V.S., Mou, H., Smits, S.L., Dekkers, D.H., Muller, M.A., Dijkman, R., Muth, D., Demmers, J.A., Zaki, A., Fouchier, R.A., et al. (2013). Dipeptidyl peptidase 4 is a functional receptor for the emerging human coronavirus-EMC. Nature 495, 251–254.

Ravenhall, M., Campino, S., Sepulveda, N., Manjurano, A., Nadjm, B., Mtove, G., Wangai, H., Maxwell, C., Olomi, R., Reyburn, H., et al. (2018). Novel genetic polymorphisms associated with severe malaria and under selective pressure in Northeastern Tanzania. PLoS Genet 14, e1007172.

Ritchie, M.E., Phipson, B., Wu, D., Hu, Y., Law, C.W., Shi, W., and Smyth, G.K. (2015). limma powers differential expression analyses for RNA-sequencing and microarray studies. Nucleic Acids Res 43, e47.

Robinson, M.D., and Oshlack, A. (2010). A scaling normalization method for differential expression analysis of RNA-seq data. Genome Biol 11, R25.

RStudio (2020). Shiny Server: Put Shiny Web Apps Online.

Scerri, T.S., Quaglieri, A., Cai, C., Zernant, J., Matsunami, N., Baird, L., Scheppke, L., Bonelli, R., Yannuzzi, L.A., Friedlander, M., et al. (2017). Genome-wide analyses identify common variants associated with macular telangiectasia type 2. Nat Genet 49, 559–567.

Seifirad, S. (2020). Pirfenidone: A novel hypothetical treatment for COVID-19. Med Hypotheses 144, 110005.

Setoh, K., Terao, C., Muro, S., Kawaguchi, T., Tabara, Y., Takahashi, M., Nakayama, T., Kosugi, S., Sekine, A., Yamada, R., et al. (2015). Three missense variants of metabolic syndrome-related genes are associated with alpha-1 antitrypsin levels. Nat Commun 6, 7754.

Shah, S., Henry, A., Roselli, C., Lin, H., Sveinbjornsson, G., Fatemifar, G., Hedman, A.K., Wilk, J.B., Morley, M.P., Chaffin, M.D., et al. (2020). Genome-wide association and Mendelian randomisation analysis provide insights into the pathogenesis of heart failure. Nat Commun 11, 163.

Sharapov, S.Z., Shadrina, A.S., Tsepilov, Y.A., Elgaeva, E.E., Tiys, E.S., Feoktistova, S.G., Zaytseva, O.O., Vuckovic, F., Cuadrat, R., Jager, S., et al. (2020). Replication of fifteen loci involved in human plasma protein N-glycosylation in 4,802 samples from four cohorts. Glycobiology.

Sharapov, S.Z., Tsepilov, Y.A., Klaric, L., Mangino, M., Thareja, G., Shadrina, A.S., Simurina, M., Dagostino, C., Dmitrieva, J., Vilaj, M., et al. (2019). Defining the genetic control of human blood plasma N-glycome using genome-wide association study. Human molecular genetics 28, 2062–2077.

Shi, H., Han, X., Jiang, N., Cao, Y., Alwalid, O., Gu, J., Fan, Y., and Zheng, C. (2020). Radiological findings from 81 patients with COVID-19 pneumonia in Wuhan, China: a descriptive study. The Lancet infectious diseases 20, 425–434.

Shima, M., Fujimura, Y., Nishiyama, T., Tsujiuchi, T., Narita, N., Matsui, T., Titani, K., Katayama, M., Yamamoto, F., and Yoshioka, A. (1995). ABO blood group genotype and plasma von Willebrand factor in normal individuals. Vox Sang 68, 236–240.

Shin, S.Y., Fauman, E.B., Petersen, A.K., Krumsiek, J., Santos, R., Huang, J., Arnold, M., Erte, I., Forgetta, V., Yang, T.P., et al. (2014). An atlas of genetic influences on human blood metabolites. Nat Genet 46, 543–550.

Sodetz, J.M., Paulson, J.C., and McKee, P.A. (1979). Carbohydrate composition and identification of blood group A, B, and H oligosaccharide structures on human Factor VIII/von Willebrand factor. J Biol Chem 254, 10754–10760.

Song, J., Chen, F., Campos, M., Bolgiano, D., Houck, K., Chambless, L.E., Wu, K.K., Folsom, A.R., Couper, D., Boerwinkle, E., et al. (2015). Quantitative Influence of ABO Blood Groups on Factor VIII and Its Ratio to von Willebrand Factor, Novel Observations from an ARIC Study of 11,673 Subjects. PLoS One 10, e0132626.

Staley, J.R., Blackshaw, J., Kamat, M.A., Ellis, S., Surendran, P., Sun, B.B., Paul, D.S., Freitag, D., Burgess, S., Danesh, J., et al. (2016). PhenoScanner: a database of human genotype-phenotype associations. Bioinformatics 32, 3207–3209.

Suhre, K., Arnold, M., Bhagwat, A.M., Cotton, R.J., Engelke, R., Raffler, J., Sarwath, H., Thareja, G., Wahl, A., DeLisle, R.K., et al. (2017). Connecting genetic risk to disease end points through the human blood plasma proteome. Nat Commun 8, 14357.

Sulem, P., Gudbjartsson, D.F., Walters, G.B., Helgadottir, H.T., Helgason, A., Gudjonsson, S.A., Zanon, C., Besenbacher, S., Bjornsdottir, G., Magnusson, O.T., et al. (2011). Identification of low-frequency variants associated with gout and serum uric acid levels. Nat Genet 43, 1127–1130.

Tanikawa, C., Urabe, Y., Matsuo, K., Kubo, M., Takahashi, A., Ito, H., Tajima, K., Kamatani, N., Nakamura, Y., and Matsuda, K. (2012). A genome-wide association study identifies two susceptibility loci for duodenal ulcer in the Japanese population. Nat Genet 44, 430-434, S431-432.

Team, R.C. (2020). R: A language and environment for statistical computing. R Foundation for Statistical Computing, Vienna, Austria. URL: http://www.R-project.org/.

Thorleifsson, G., Holm, H., Edvardsson, V., Walters, G.B., Styrkarsdottir, U., Gudbjartsson, D.F., Sulem, P., Halldorsson, B.V., de Vegt, F., d’Ancona, F.C., et al. (2009). Sequence variants in the CLDN14 gene associate with kidney stones and bone mineral density. Nat Genet 41, 926–930.

Timmann, C., Thye, T., Vens, M., Evans, J., May, J., Ehmen, C., Sievertsen, J., Muntau, B., Ruge, G., Loag, W., et al. (2012). Genome-wide association study indicates two novel resistance loci for severe malaria. Nature 489, 443–446.

Tin, A., Marten, J., Halperin Kuhns, V.L., Li, Y., Wuttke, M., Kirsten, H., Sieber, K.B., Qiu, C., Gorski, M., Yu, Z., et al. (2019). Target genes, variants, tissues and transcriptional pathways influencing human serum urate levels. Nat Genet 51, 1459–1474.

Tin, A., Woodward, O.M., Kao, W.H., Liu, C.T., Lu, X., Nalls, M.A., Shriner, D., Semmo, M., Akylbekova, E.L., Wyatt, S.B., et al. (2011). Genome-wide association study for serum urate concentrations and gout among African Americans identifies genomic risk loci and a novel URAT1 loss-of-function allele. Human molecular genetics 20, 4056–4068.

Visscher, P.M., Wray, N.R., Zhang, Q., Sklar, P., McCarthy, M.I., Brown, M.A., and Yang, J. (2017). 10 Years of GWAS Discovery: Biology, Function, and Translation. Am J Hum Genet 101, 5–22.

Wang, L., Oehlers, S.H., Espenschied, S.T., Rawls, J.F., Tobin, D.M., and Ko, D.C. (2015). CPAG: software for leveraging pleiotropy in GWAS to reveal similarity between human traits links plasma fatty acids and intestinal inflammation. Genome Biol 16, 190.

Wang, L., Pittman, K.J., Barker, J.R., Salinas, R.E., Stanaway, I.B., Williams, G.D., Carroll, R.J., Balmat, T., Ingham, A., Gopalakrishnan, A.M., et al. (2018). An Atlas of Genetic Variation Linking Pathogen-Induced Cellular Traits to Human Disease. Cell Host Microbe 24, 308–323 e306.

Welter, D., MacArthur, J., Morales, J., Burdett, T., Hall, P., Junkins, H., Klemm, A., Flicek, P., Manolio, T., Hindorff, L., et al. (2014). The NHGRI GWAS Catalog, a curated resource of SNP-trait associations. Nucleic Acids Res 42, D1001–1006.

Wool, G.D., and Miller, J.L. (2020). The Impact of COVID-19 Disease on Platelets and Coagulation. Pathobiology, 1-13.

Zhao, J., Yang, Y., Huang, H., Li, D., Gu, D., Lu, X., Zhang, Z., Liu, L., Liu, T., Liu, Y., et al. (2020). Relationship between the ABO Blood Group and the COVID-19 Susceptibility. Clin Infect Dis.

Zhong, F.L., Robinson, K., Teo, D.E.T., Tan, K.Y., Lim, C., Harapas, C.R., Yu, C.H., Xie, W.H., Sobota, R.M., Au, V.B., et al. (2018). Human DPP9 represses NLRP1 inflammasome and protects against autoinflammatory diseases via both peptidase activity and FIIND domain binding. J Biol Chem 293, 18864–18878.

Zhu, Z., Anttila, V., Smoller, J.W., and Lee, P.H. (2018). Statistical power and utility of meta-analysis methods for cross-phenotype genome-wide association studies. PLoS One 13, e0193256.

