## SupplementalFiguresandTables for "An atlas connecting shared genetic architecture of human diseases and molecular phenotypes provides insight into COVID-19 susceptibility"

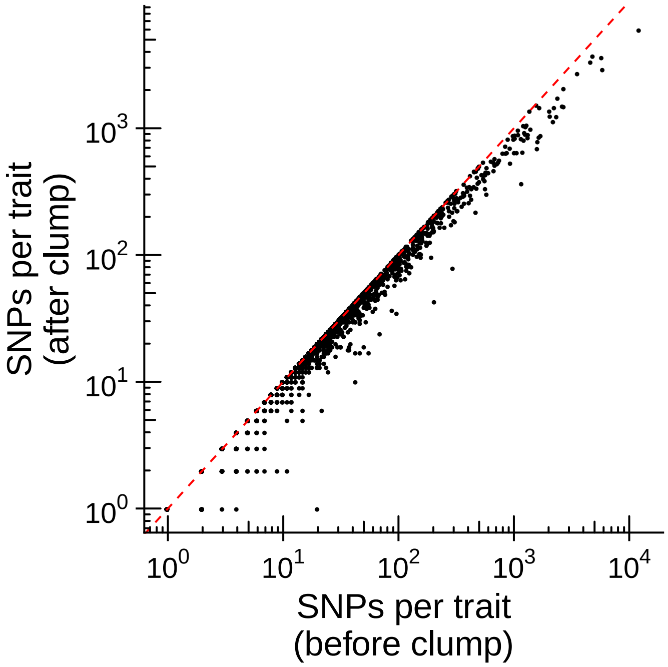


**Figure S1.** Related to Figure 1. Clumping of GWAS results from NHGRI-EBI GWAS catalog. For each GWAS traits, we performed LD clumping to only keep the lead SNP in each region.

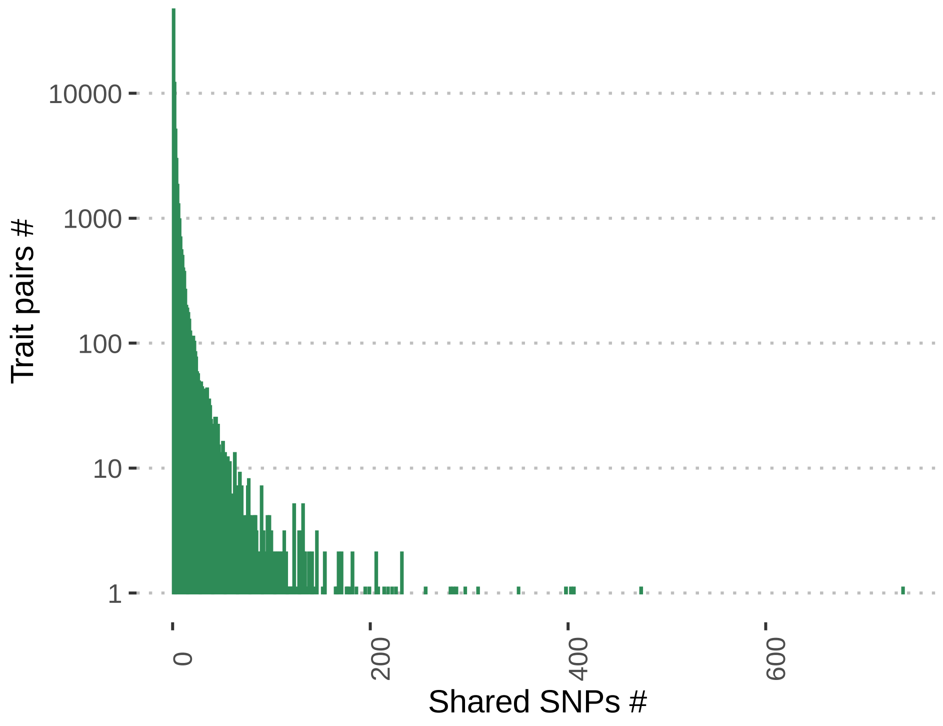


**Figure S2**. Histogram of shared SNPs for each trait pair in NHGRI-EBI GWAS catalog from iCPAGdb at false discovery rate of 0.1. Among 76127 trait pairs (including compound phenotypes), the mean number of shared independent SNPs was 3.10, and 38.8% of trait pairs shared more than 1 SNP. “Mathematical ability” and “self reported educational attainment” shared the most SNPs with 739.

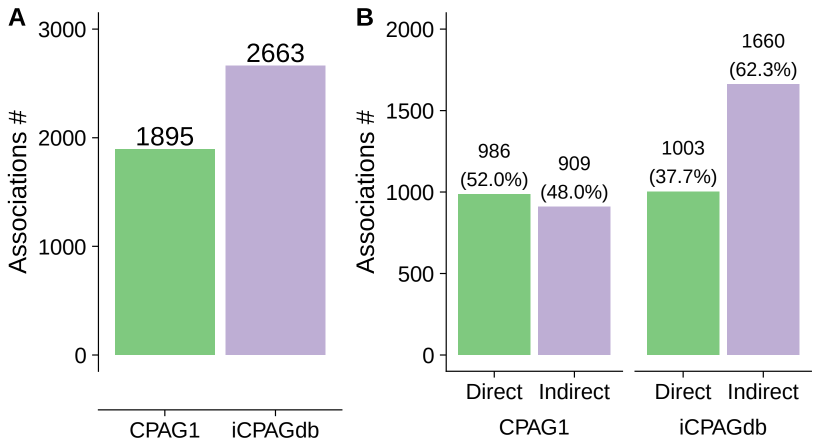


**Figure S3.** Related to Figure 1. Comparison of CPAG1 and iCPAGdb using the NHGRI-EBI GWAS catalog summary statistics downloaded on September 4, 2013. CPAG1 finished this analysis in 73.2 minutes while iCPAGdb used 4.1 minutes on a MacBook with Quad-Core intel i7 CPU and 16 GBs of RAM. CPAG1 cannot run efficiently enough to allow for comparison with the 2020 NHGRI-EBI GWAS catalog (estimated completion time > ~10 days for CPAG vs. ~100 minutes for iCPAGdb). A total of 887 traits were included in the dataset, and p values were corrected using multiple test correction for 392,941 comparisons. A) iCPAGdb detected 2598 cross-phenotype associations, 37% more than CPAG1 at FDR of 0.1. B) The number of cross-phenotype associations from directly shared SNPs were nearly unchanged between iCPAGdb and CPAG1 but indirect associations increased 76%.

**Table S1.** Related to Figure 1. iCPAGdb output for cross-phenotype associations from NHGRI-EBI GWAS catalog (downloaded on August 5, 2020).

**Table S2.** Related to Figure 2. iCPAGdb output for cross-phenotype associations between molecular and cellular datasets and NHGRI-EBI GWAS catalog.

**Table S3.** Related to Figure 2. COLOC analysis of *CXCL10* following *C. trachomatis* infection and levels of *CXCL9* (MIG) in whole blood.

**Table S4.** Related to Figure 3. Cross-phenotype associations for COVID-19 (p value < $1 \times 10^{-5}$) against 4414 GWAS traits in iCPAGdb (p value < $5 \times 10^{-8}$).

**Table S5.** Related to Figure 3. COLOC analysis output for *ABO* region between COVID-19 and CD209 antigen levels.

**Table S6.** Related to Figure 4. COLOC analysis output for *DPP9* region between COVID-19 and idiopathic pulmonary fibrosis.
