## SupplementalNote_iCPAGdbWebApp for "An atlas connecting shared genetic architecture of human diseases and molecular phenotypes provides insight into COVID-19 susceptibility"

### iCPAGdb Web App Supplement

Thomas J. Balmat, Research Computing, Duke University

Two primary objectives guided the design of the iCPAGdb web app (the web app): quick accessibility of precomputed cross-phenotype analysis of GWAS (CPAG) results for select GWAS studies and the ability to discover new cross-phenotype relationships between researcher uploaded GWAS data sets and those maintained in the iCPAGdb database (the database). We will refer to the browsing and upload modes simply as "review" and "compute," respectively. Once the data content and public interaction objectives were established, architecture design choices were made, drawing on experience gained while developing a web app for a prior GWAS project (Wang et al., 2018). Primary software components for the web portion of iCPAGdb are the R statistical programming language (R Core Team, 2020), the R package *Shiny* for interaction of web pages with R scripts (Chang et al., 2020), *Shiny Server* as a 24/7 multi-user platform to make Shiny apps publicly accessible (RStudio, 2020), the database environment *SQLite* for efficient querying of GWAS and CPAG results (Hipp, 2020), and the R package *RSQLite* to execute SQL queries from within R scripts (Muller et al., 2020). CPAG computations, as described in the iCPAGdb section of the paper and (Wang et al., 2015), are implemented in the Python programming language, external to the web app, and are executed using parameterized system calls constructed from values supplied by the user through on-screen controls. The results of a CPAG execution are read by the R script, processed, and presented to the viewer in various tables and graphs on a web page. Shiny conducts the interaction between web pages, R scripts, and the CPAG functions. The SQLite database is primarily used by the CPAG functions, but is also a source of identifiers and other values used to populate selection lists within the web app.

Program flow 1 outlines the major processing steps for review mode. Basically, the user requests an existing CPAG result set from which a corresponding table and heatmap are generated and displayed. Various filtering and graph construction controls are available for iterative sub-setting of data and selection of significance measure and number of top significant phenotype pairs to plot. The "Download" button enables the researcher to make a local copy of records appearing in the currently displayed results table. Important packages used in this mode are DT (Xie et al., 2020) for construction of and interaction with tables and ggplot2 (Wickham, 2016), plotly (Sievert, 2020), and heatmaply (Galili et al., 2017) for basic plotting, interactive plotting (hover labels), and heatmap generation, respectively.

Program flow 1, review mode

1. Query available precomputed CPAG results and construct table with selectable rows, one per study
2. Present table in web app
3. When table row is selected, read corresponding study results, prepare table of results and heatmap for review
4. Interact with user, iterating through on-screen control settings to sort, filter, and modify results table while synchronizing heatmap with tabular data

Although the packages employed are relatively full-featured and robust, several custom algorithms had to be developed to overcome specific limitations of various functions of these packages when used with our data. For example, the clustering algorithm used by the heatmaply() function to add dendrogram lines to a graph generated errors when used with the distance matrix computed by the base R distance() function. A substitute vector distance algorithm was developed to overcome domain errors reported by heatmaply(). Figure 1 shows an example review session.[1] The table in the upper section of the form lists precomputed CPAG results available for review. Below that are the filter and graph controls. When a row in the CPAG table is selected, a table of corresponding results appears in the "Table" tab (bottom of the image) and a corresponding heatmap that relates pairs of inter-GWAS phenotypes by the selected significance measure (Fisher, Bonferroni, etc.) appears in the "Heatmap" tab. Figures 2 and 3 show the table and heatmap that appear after clicking the "Molecular traits vs. Human disease" row of the selection table, which is accessed by scrolling down.

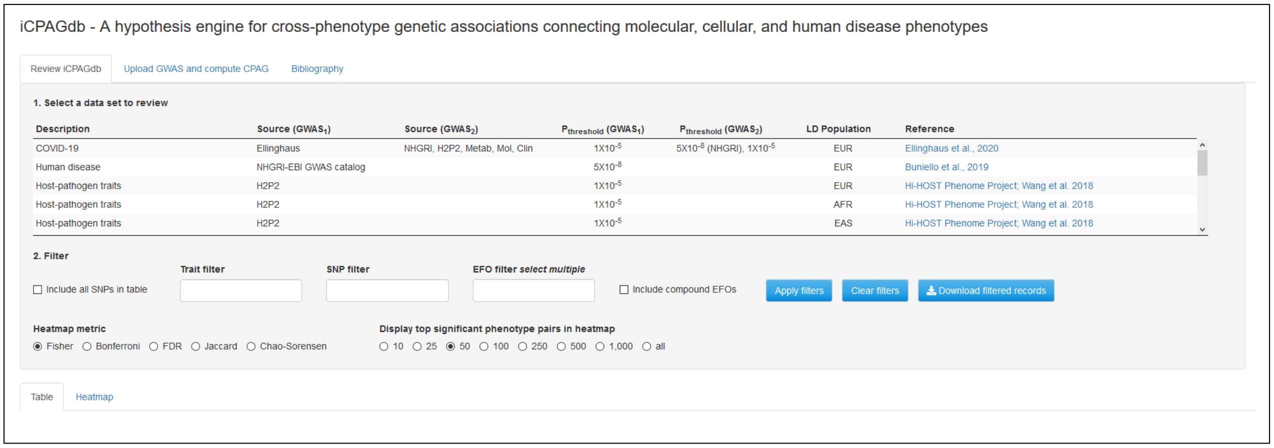


Figure 1: iCPAGdb web app Review tab

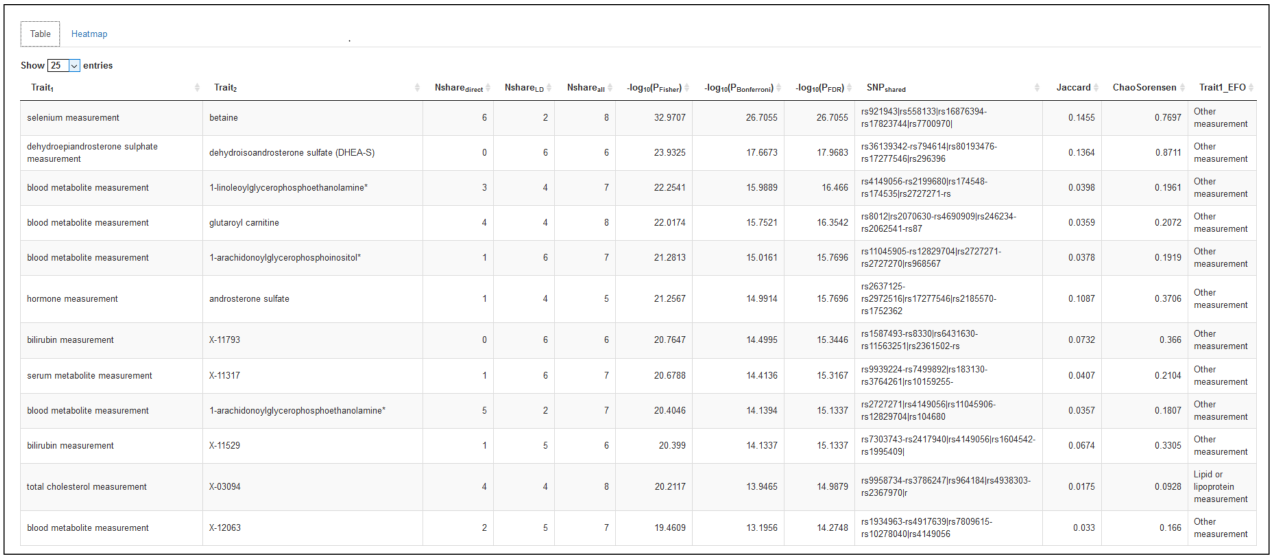

Table | Heatmap

Show 25 entries

| $Trait_1$ | $Trait_2$ | $Nshare_{direct}$ | $Nshare_{LD}$ | $Nshare_{all}$ | $-log_{10}(P_{Fisher})$ | $-log_{10}(P_{Bonferroni})$ | $-log_{10}(P_{FDR})$ | $SNP_{shared}$ | Jaccard | ChaoSorensen | Trait1_EFO |
|---|---|---|---|---|---|---|---|---|---|---|---|
| selenium measurement | betaine | 6 | 2 | 8 | 32.9707 | 26.7055 | 26.7055 | rs921943\|rs558133\|rs16876394-rs17823744\|rs7700970\| | 0.1455 | 0.7697 | Other measurement |
| dehydroepiandrosterone sulphate measurement | dehydroisoandrosterone sulfate (DHEA-S) | 0 | 6 | 6 | 23.9325 | 17.6673 | 17.9683 | rs36139342-rs794614\|rs80193476-rs17277546\|rs296396 | 0.1364 | 0.8711 | Other measurement |
| blood metabolite measurement | 1-linoleoylglycerophosphoethanolamine* | 3 | 4 | 7 | 22.2541 | 15.9889 | 16.466 | rs4149056-rs2199680\|rs174548-rs174535\|rs2727271-rs | 0.0398 | 0.1961 | Other measurement |
| blood metabolite measurement | glutaroyl carnitine | 4 | 4 | 8 | 22.0174 | 15.7521 | 16.3542 | rs8012\|rs2070630-rs4690909\|rs246234-rs2062541-rs87 | 0.0359 | 0.2072 | Other measurement |
| blood metabolite measurement | 1-arachidonoylglycerophosphoinositol* | 1 | 6 | 7 | 21.2813 | 15.0161 | 15.7696 | rs11045905-rs12829704\|rs2727271-rs2727270\|rs968567 | 0.0378 | 0.1919 | Other measurement |
| hormone measurement | androsterone sulfate | 1 | 4 | 5 | 21.2567 | 14.9914 | 15.7696 | rs2637125-rs2972516\|rs17277546\|rs2185570-rs1752362 | 0.1087 | 0.3706 | Other measurement |
| bilirubin measurement | X-11793 | 0 | 6 | 6 | 20.7647 | 14.4995 | 15.3446 | rs1587493-rs8330\|rs6431630-rs11563251\|rs2361502-rs | 0.0732 | 0.366 | Other measurement |
| serum metabolite measurement | X-11317 | 1 | 6 | 7 | 20.6788 | 14.4136 | 15.3167 | rs9939224-rs7499892\|rs183130-rs3764261\|rs10159255- | 0.0407 | 0.2104 | Other measurement |
| blood metabolite measurement | 1-arachidonoylglycerophosphoethanolamine* | 5 | 2 | 7 | 20.4046 | 14.1394 | 15.1337 | rs2727271\|rs4149056\|rs11045906-rs12829704\|rs104680 | 0.0357 | 0.1807 | Other measurement |
| bilirubin measurement | X-11529 | 1 | 5 | 6 | 20.399 | 14.1337 | 15.1337 | rs7303743-rs2417940\|rs4149056\|rs1604542-rs1995409\| | 0.0674 | 0.3305 | Other measurement |
| total cholesterol measurement | X-03094 | 4 | 4 | 8 | 20.2117 | 13.9465 | 14.9879 | rs9958734-rs3786247\|rs964184\|rs4938303-rs2367970\|r | 0.0175 | 0.0928 | Lipid or lipoprotein measurement |
| blood metabolite measurement | X-12063 | 2 | 5 | 7 | 19.4609 | 13.1956 | 14.2748 | rs1934963-rs4917639\|rs7809615-rs10278040\|rs4149056 | 0.033 | 0.166 | Other measurement |

Figure 2: iCPAGdb web app. Table generated by selecting a precomputed CPAG result.

[1]Screen-shots were taken from the iCPAGdb website, http://cpag.oit.duke.edu

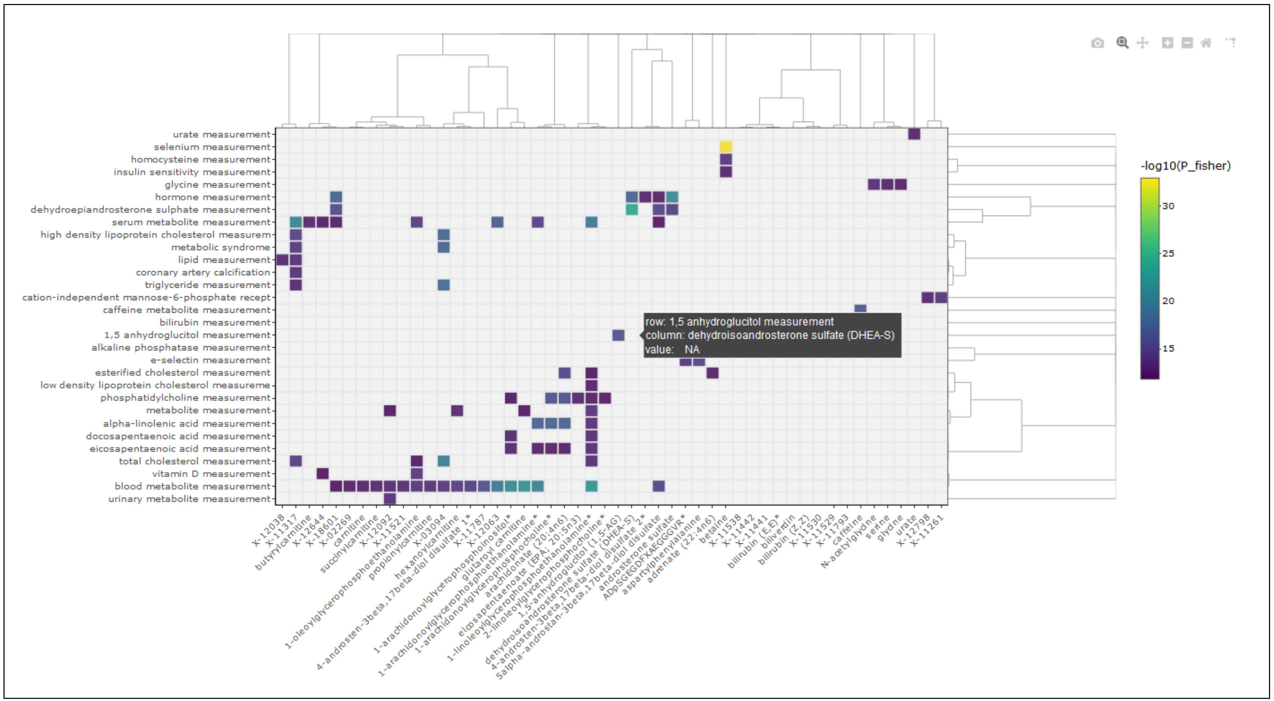


Figure 3: iCPAGdb web app. Heatmap generated by selecting a precomputed CPAG result.

Program flow 2 outlines the major processing steps for compute mode. Figure 4 shows the corresponding “Upload and Compute” tab of the app. In this mode, the user browses files on a local computer, selects a properly formatted GWAS result file of interest (containing, for a single phenotype, SNPs and GWAS significance, p, values), specifies format and column configuration, then uploads the file. Next, CPAG computation parameter values, including iCPAGdb GWAS set to be crossed with, significance thresholds for filtering, and linkage disequilibrium (LD) population are specified. When “Compute CPAG” is pressed, the R script composes a system level command to execute the CPAG (Python) function. The future() function of the R future package (Bengtsson, 2020) combined with a delaying pipe from the promises package (Cheng, 2020) execute CPAG operations asynchronously, waiting on completion before resuming R script execution. An important consideration is that the default behavior of R is to execute instructions synchronously, so that one instruction is completed in entirety prior to another beginning. This is problematic in a multi-user setting when long-running computations are executed. Typical CPAG execution time ranges from thirty seconds to ten minutes, making synchronous execution problematic. Although (the open source version of) Shiny Server accommodates multiple simultaneous connections, it allocates a single R process to each application. iCPAGdb is an application. Therefore, within the group of simultaneous iCPAGdb users, at most one is being serviced at any given time. Others must wait until that user’s CPAG analysis completes. But, by executing the CPAG function from within a future() call and promise pipe sequence, R will spawn an individual, asynchronous process for executing the CPAG function, enabling multiple, simultaneous users. Of course, this has to be programmed and scripts must be adjusted to account for the delayed, and impromptu nature of results being returned. In addition to one CPU being allocated for each asynchronous CPAG call, the CPAG function itself executes as a multi-threaded process on multiple CPUs. It is important to consider expected load (number of simultaneous users and number of CPUs used in parallel) when configuring a server to be used for public compute services.

Program flow 2, compute mode

**Upload a GWAS section**

Researcher:

1. Browses local computer for a GWAS file and uploads it
2. Specifies file structure (delimiter type, GWAS SNP and significance columns)

**Compute section**

Researcher:

3. Selects iCPAGdb GWAS study (GWAS source two) to be used for investigation of cross-GWAS associations
4. Selects p-thresholds (to filter phenotypes and SNPs) for each GWAS and specifies a linkage disequilibrium (LD) population
5. Clicks "Compute CPAG"

The R script:

6. Validates the uploaded file (verifies that specified columns and delimiters are present)
7. Composes a CPAG (Python) function call, using researcher supplied values
8. Creates an asynchronous future() environment for CPAG execution
9. Passes control to the CPAG function and waits on returned (promise piped) results
10. Generates a table of results and heatmap using current on-screen configuration values

Researcher:

11. Reviews and interacts with results as on the Review tab

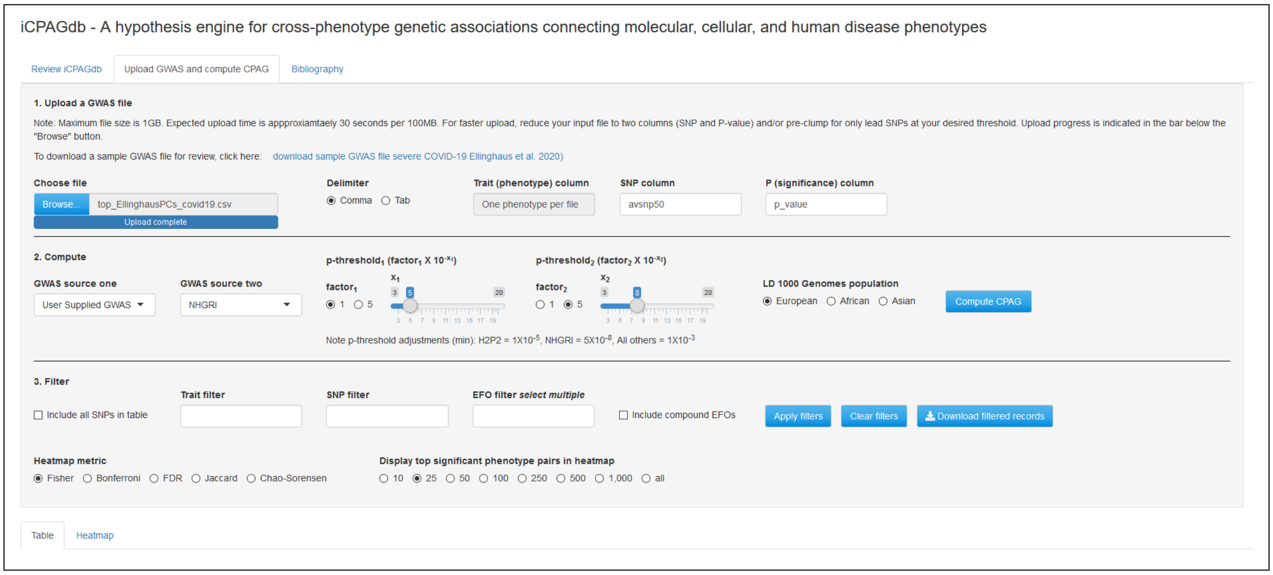


Figure 4: iCPAGdb web app. Upload and Compute tab..

In initial testing, the app was found to perform efficiently, giving researchers convenient access to both precomputed CPAG results and newly computed values for uploaded GWAS data. Cross-phenotype relationships presented by the app agree with those known to researchers with expert knowledge of data sets employed during verification. It is hoped that the app will become a recognized and useful tool for researchers conducting exploratory cross-phenotype analysis of GWAS.

Complete R and Shiny scripts for the iCPAGdb web app, along with additional design information, are available at https://github.com/tbalmat/iCPAGdb.

### References


Henrik Bengtsson. A unifying framework for parallel and distributed processing in r using futures, aug 2020. URL `https://arxiv.org/abs/2008.00553`.

Winston Chang, Joe Cheng, JJ Allaire, Yihui Xie, and Jonathan McPherson. *Shiny: Web Application Framework for R*, 2020. URL `https://CRAN.R-project.org/package=shiny`. R package version 1.5.0.

Joe Cheng. *promises: Abstractions for Promise-Based Asynchronous Programming*, 2020. URL `https://CRAN.R-project.org/package=promises`. R package version 1.1.1.

Galili, Tal, O'Callaghan, Alan, Sidi, Jonathan, Sievert, and Carson. heatmaply: an r package for creating interactive cluster heatmaps for online publishing. *Bioinformatics*, 2017. doi: 10.1093/bioinformatics/btx657. URL `http://dx.doi.org/10.1093/bioinformatics/btx657`.

Richard D Hipp. *SQLite*, 2020. URL `https://www.sqlite.org/index.html`.

Kirill Muller, Hadley Wickham, David A. James, and Seth Falcon. *RSQLite: 'SQLite' Interface for R*, 2020. URL `https://CRAN.R-project.org/package=RSQLite`. R package version 2.2.1.

R Core Team. *R: A Language and Environment for Statistical Computing*. R Foundation for Statistical Computing, Vienna, Austria, 2020. URL `https://www.R-project.org/`.

RStudio. *Shiny Server: Put Shiny Web Apps Online*. RStudio, 2020. URL `https://rstudio.com/products/shiny/shiny-server/`. Version 1.5.0.

Carson Sievert. *Interactive Web-Based Data Visualization with R, plotly, and shiny*. Chapman and Hall/CRC, 2020. ISBN 9781138331457. URL `https://plotly-r.com`.

L. Wang, S.H. Oehlers, S.T. Espenschied, J.F. Rawls, D.M. Tobin, and D.C. Ko. CPAG: software for leveraging pleiotropy in gwas to reveal similarity between human traits links plasma fatty acids and intestinal inflammation. *Genome Biology*, 16, 9 2015. URL `https://genomebiology.biomedcentral.com/articles/10.1186/s13059-015-0722-1`.

L. Wang, K.J. Pittman, J.R. Barker, R.E. Salinas, I.B. Stanaway, R.J. Williams, G.D. ad Carroll, T. Balmat, A. Ingham, and et al. Gopalakrishnan, A.M. An atlas of genetic variation linking pathogen-induced cellular traits to human disease. *Cell & Host Microbe*, 24:308–323, 8 2018. URL `https://www.cell.com/cell-host-microbe/fulltext/S1931-3128(18)30377-9`.

Hadley Wickham. *ggplot2: Elegant Graphics for Data Analysis*. Springer-Verlag New York, 2016. ISBN 978-3-319-24277-4. URL `https://ggplot2.tidyverse.org`.

Yihui Xie, Joe Cheng, and Xianying Tan. *DT: A Wrapper of the JavaScript Library 'DataTables'*, 2020. URL `https://CRAN.R-project.org/package=DT`. R package version 0.16.
